# Triangulating evidence from the UK Biobank and China shows the health and behavioral impact of vegetarianism

**DOI:** 10.1101/2025.03.09.25323635

**Authors:** Chen Zhu, Xiaosong Yang, Yanjun Song, Wenyan Xu, Jiakai Gong, Xiaolu Wang, Wei Si, Shenggen Fan

## Abstract

**Background:** Vegetarianism is widely recognized for its health and environmental benefits. However, its broader impacts on physical, mental, and social well-being remain underexplored. This study investigates the health and behavioral outcomes associated with vegetarianism across diverse populations.

**Methods:** We analysed polygenic scores for vegetarianism (VegPGS) in 495,971 UK Biobank (UKB) participants and performed phenome-wide association studies (PheWAS) on 443 health and behavioral traits. Cross-validation analyses were conducted using data from 9,009 vegetarians and 486,962 non-vegetarians. One- and two-sample Mendelian randomization (MR) analyses explored causal relationships. Findings were further validated in 11,642 participants from the China Health and Nutrition Survey (CHNS). Additionally, machine-learning classification models were developed to predict vegetarian status using behavioral, physiological, and genetic factors.

**Findings:** PheWAS identified 57 health-related and 1 behavior-related factor significantly associated with VegPGS, with cross-validation confirming these links. MR analyses supported causal effects of vegetarianism on lower basal metabolic rate, reduced body mass index (BMI), decreased fat mass, and lower risk of type 2 diabetes. CHNS data confirmed associations with lower BMI and diabetes risk in East Asian populations. Machine-learning models achieved high accuracy in predicting vegetarian status (AUC 0.913C±C0.018).

**Interpretation:** This study provides robust evidence supporting the metabolic health benefits of vegetarianism. The integration of multimodal genetic, behavioral, and physiological data enhances understanding and prediction of dietary choices, offering valuable insights for policymakers and individuals considering a transition to plant-based diets to achieve sustainability.

**Funding:** National Natural Science Foundation of China (Nos. 72103187 and 72061147002) and the 2115 Talent Development Program at China Agricultural University.

## 1. Introduction

Vegetarianism is a dietary pattern associated with significant benefits for human health and environmental sustainability.[1–2] A plant-based diet is linked to reduced risks of chronic diseases, including cardiovascular conditions and some cancers, while also contributing to lower greenhouse gas (GHG) emissions and sustainable resource use.[3] However, the broader impact of vegetarianism extends beyond these health outcomes, affecting various aspects of physical, mental, and social well-being. Existing research on vegetarianism has largely focused on specific health outcomes, such as reduced risks of cardiovascular disease (CVD), diabetes, and colorectal cancer, but it has also reported potential adverse effects, such as the link with an increased risk of depression.[4–8] While these studies provide valuable insights, they often rely on observational methods that are prone to confounders and may lack the rigour required to establish causal relationships.[8] Moreover, the effects of vegetarianism on psychological well-being, social interactions, and micronutrient deficiencies remain underexplored, which limits our understanding of the true breadth of vegetarianism’s impact.

A more comprehensive exploration that integrates causal inference and considers a broader set of health and behavioral factors is needed to deepen our understanding of vegetarianism. By broadening the scope, researchers can identify both positive and potentially adverse effects, providing a clearer, more nuanced perspective on how a vegetarian diet affects individual and public health. Additionally, incorporating economic and behavioral theories could offer valuable insights into the decision-making processes that underlie dietary choices related to vegetarianism. For instance, the random utility theory (RUT) explains individual choice by modelling decision-making as a process where individuals choose the option that maximizes their perceived utility or satisfaction.[9] In this theoretical framework, utility can be first constructed by demographic, socioeconomic, behavioral, physiological, genetic, and even multi-omic factors, and then used to predict individual choices. However, applications on individuals’ decision-making behaviour related to dietary choices have been scarce.[10]

This study aims to address these gaps by systematically investigating a wide array of health, physiological, and behavior-related outcomes associated with vegetarianism. The goal is to provide a holistic understanding of the implications of vegetarian diets, offering insights for nutritionists, policymakers, and individuals considering a transition to plant-based diets to achieve sustainability. The analysis is grounded in robust causal inference and machine-learning methodologies, addressing limitations of prior observational studies.

Using data from the UK Biobank (UKB; N = 495,971), we conducted a phenome-wide association study (PheWAS) to explore the relationships between genetic predisposition for vegetarianism—captured through polygenic scores (VegPGS)—and 443 health/physiological and behavioral traits. We validated these associations at the phenotypic level using multivariate linear and logistic regression analyses. Further, we applied both one-sample and two-sample Mendelian randomization (MR) to investigate potential causal relationships between vegetarianism and the identified health and behavior-related outcomes. To enhance the generalizability of our findings, we examined significant causal associations in East Asian populations using an independent cohort from the China Health and Nutrition Survey (CHNS; N = 11,642). Finally, to better understand the decision-making processes underlying individual dietary choices, we developed machine-learning classification models incorporating behavioral, physiological, and genetic factors based on the random utility theory to improve the prediction accuracy of individual dietary choices related to vegetarianism. Fig. 1 provides an overview of the study design.

**Fig. 1.**
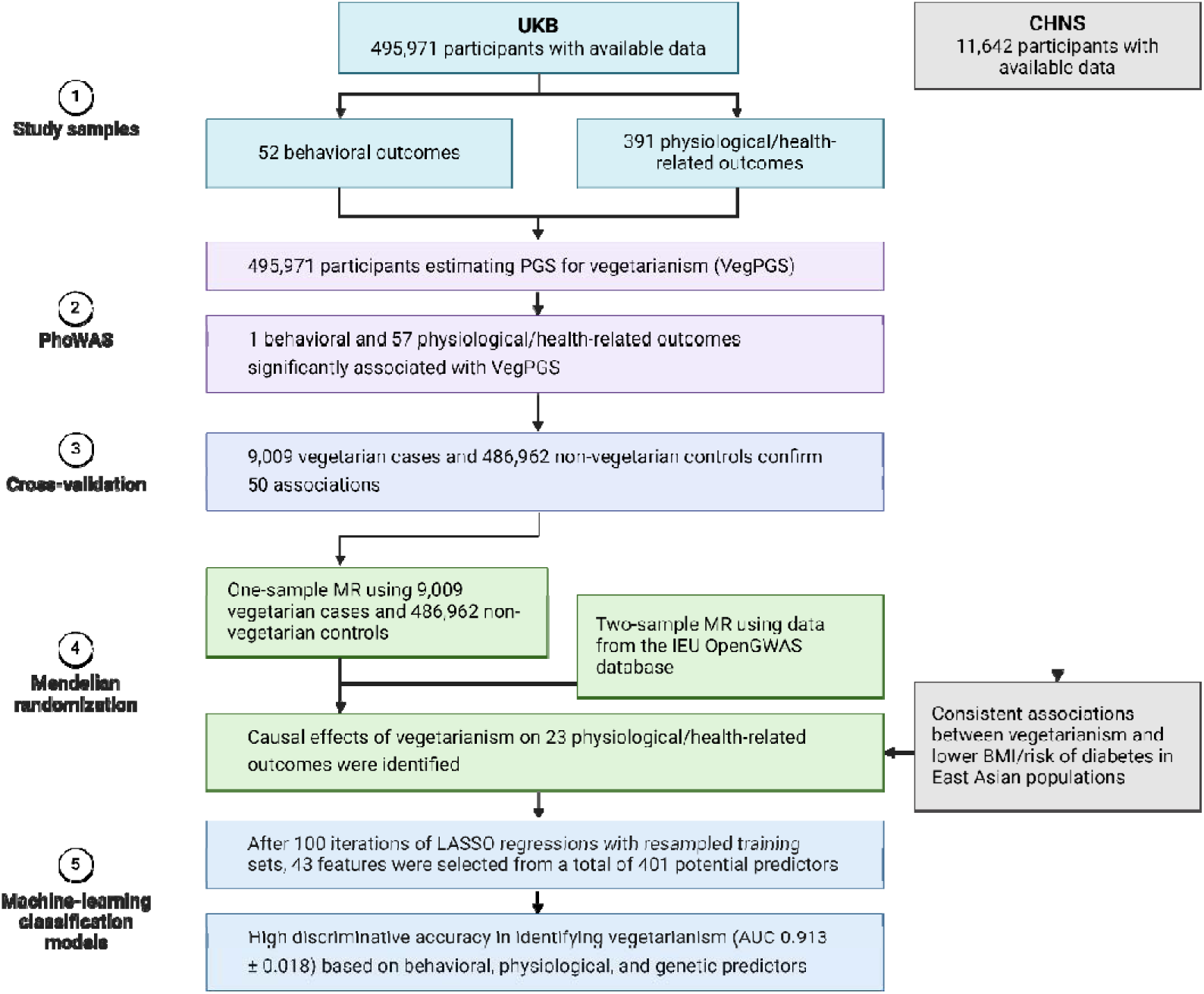
Overview of the study design. Procedures to identify health and behavioral impacts of vegetarianism in UKB and CHNS.

## 2. Methods

### Study samples

Two independent large cohorts from the UK Biobank (UKB) and the China Health and Nutrition Survey (CHNS) databases were used in the current study. UK Biobank is the database for a population-based study involving more than 500,000 UK residents approved by the NHS National Research Ethics Service (Ref: 11/NW/0382).[11] Between March 2006 and July 2010, individuals residing within 25 miles of one of the 22 study assessment centers in England, Scotland, and Wales were recruited to provide data on a wide range of socio-demographic, clinical, and lifestyle outcomes. Blood, urine, and saliva samples, as well as physical measurements, were collected from all participants with their written informed consent during the interviews. At the baseline (2006), a food frequency touchscreen questionnaire was used to collect data on all participants’ dietary habits, including single food intakes, food types, and intake frequencies. Observations (7,848 participants) with unrealistic answers were dropped (495,971 participants remained).[12–16]

CHNS is an ongoing large-scale, longitudinal, household-based survey initiated in 1989.[17] It enrolled more than 30,000 Chinese residents in nine provinces and municipal cities using multistage random-cluster sampling methods, with 85% being Han Chinese and 15% being other East Asian ethnic groups.[18] Dietary intake was assessed in 1997, 2000, 2004, 2006, 2009, and 2011. Ethical approval for CHNS was granted by the institutional review committees of the University of North Carolina at Chapel Hill and National Institute for Nutrition and Health, Chinese Center for Disease Control and Prevention. Participants provided their written, informed consent in the survey. Additional details can be found at https://www.cpc.unc.edu/projects/china. Participants with valid dietary assessment in 2004 and at least one valid health assessment in 2006–2011 were included in the analysis (11,642 participants remained).

### Measurement of vegetarianism

A binary variable of vegetarian is defined based on participants’ profiles of food intake. Specifically, individuals who did not report any intake of processed meat, beef, pork, lamb/mutton, poultry, fish, seafood and other meat were classified as vegetarians. This resulted in 9,009 vegetarians/486,962 non-vegetarians in the analytical UKB sample, and 2,658 vegetarians/8,984 non-vegetarians in the analytical CHNS sample.

### Polygenic score for vegetarianism (VegPGS)

Genotyping, imputation, and quality control of the genetic data were performed by the UK Biobank team (http://www.ukbiobank.ac.uk/scientists-3/genetic-data/). Polygenetic score (PGS) for vegetarianism was calculated using a weighted method: PRS = (β_1_ × SNP_1_ + β_2_ × SNP_2_ + … + β_n_ × SNP_n_) × (n/sum of the β coefficients). Each SNP was assigned a code of 0, 1, or 2 based on the number of risk alleles identified. The β coefficient values were obtained from the published GWAS analysis.[19] The calculated PGS for vegetarianism had a mean of -0.572 and a standard deviation of 0.739, in which higher scores indicating a greater genetic predisposition of vegetarianism.

### Health-related phenotypes

A total of 391 physiological and health-related outcomes from five categories were analyzed in the UKB sample, including (1) 136 diseases, (2) 10 blood chemistry biomarkers, (3) 44 physical measures, (4) 4 telomere phenotypes, and (5) 197 nuclear magnetic resonance (NMR) metabolomic biomarkers. More details on health and subsequent behavior-related phenotypes were provided in Supplementary Table 1.

### Behavior-related phenotypes

Behavior-related phenotypes consist of six categories, containing a total of 52 traits: (1) 9 socio-demographic factors (such as education, income, and employment); (2) 25 lifestyle factors (such as alcohol drinking status, social activities, and outdoor activities); (3) 13 mental well-being phenotypes; (4) 5 early-life factors.

### Statistical analyses

#### PheWAS

We conducted a phenome-wide association study to explore associations between the genetic predisposition for vegetarianism, as captured by the polygenic score (VegPGS), and a total of 443 phenotypes available in the UK Biobank dataset. All analyses were performed using R (version 4.4.1). To account for potential confounders, all association tests were adjusted for sex, age, and the first ten genetic principal components to control for population stratification. In each regression, we excluded those with missing data on particular phenotypes, resulting in 28,582 to 458,709 participants in different regressions (Table 1a). Two-sided statistical tests were used for all analyses, and significance was determined using a Bonferroni correction to adjust for multiple comparisons across the 443 tests, maintaining a significance level of α = 0.05. This correction helped minimize the risk of Type I errors due to multiple hypothesis testing.

**Table 1.** Summary information.

a. Summary of phenotype groups in PheWAS analyses
| Type | Group | Number of traits | Sample size (mean [range]) |
| --- | --- | --- | --- |
| Behavior-related outcomes | Sociodemographics | 9 | 406,338 [258,961-458,709] |
| Behavior-related outcomes | Early life factors | 5 | 421,136 [350,968-457,524] |
| Behavior-related outcomes | Lifestyle factors | 25 | 394,695 [168,592-458,709] |
| Behavior-related outcomes | Mental well-being | 13 | 347,800 [109,621-458,709] |
| Physiological and health-related outcomes | Physical measures | 44 | 375,613 [28,582-457,195] |
| Physiological and health-related outcomes | Telomere length | 4 | 333,289 [1,282-443,958] |
| Physiological and health-related outcomes | Blood biochemistry | 10 | 432,834 [400,155-445,128] |
| Physiological and health-related outcomes | Diseases outcomes | 136 | 458,709 [458,709-458,709] |
| Physiological and health-related outcomes | NMR metabolomics | 197 | 256,324 [245,659-256,704] |

b. Summary statistics of vegetarian and non-vegetarian groups in UKB sample
| Characteristics | Pooled<br>(N = 495,971) | Vegetarians<br>(N = 9,009) | Non-vegetarians<br>(N = 486,962) |
| --- | --- | --- | --- |
| Age, mean (SD) | 56.5 (8.1) | 53.2 (7.9) | 56.6 (8.1) |
| Sex, N (%) |  |  |  |
| *Female* | 269,883 (54.4%) | 5,995 (66.5%) | 263,888 (54.2%) |
| *Male* | 226,088 (45.6%) | 3,014 (33.5%) | 223,074 (45.8%) |
| College, N (%) | 160,879 (32.4%) | 4,470 (49.6%) | 156,409 (32.1%) |
| BMI, mean (SD) | 27.4 (4.8) | 25.7 (4.7) | 27.5 (4.8) |
| Region, N (%) |  |  |  |
| *North East* | 55,647 (11.2%) | 658 (7.3%) | 55,026 (11.3%) |
| *North West* | 77,818 (15.7%) | 1,157 (12.8%) | 76,657 (15.7%) |
| *Yorkshire and The Humber* | 69,833 (14.1%) | 1,094 (12.1%) | 68,714 (14.1%) |
| *East Midlands* | 40,025 (8.1%) | 790 (8.8%) | 39,252 (8.1%) |
| *West Midlands* | 40,868 (8.2%) | 894 (9.9%) | 39,984 (8.2%) |
| *East of England* | 1,438 (0.3%) | 38 (0.4%) | 1,371 (0.3%) |
| *London* | 61,798 (12.5%) | 1,994 (22.1%) | 59,803 (12.3%) |
| *South East* | 48,853 (9.9%) | 824 (9.2%) | 48,019 (9.9%) |
| *South West* | 41,910 (8.5%) | 744 (8.3%) | 41,153 (8.5%) |
| *Wales* | 21,476 (4.3%) | 358 (4.0%) | 21,138 (4.3%) |
| *Scotland* | 36,305 (7.3%) | 458 (5.1%) | 35,845 (7.4%) |

c. Summary statistics of vegetarian and non-vegetarian groups in CHNS sample
| Characteristics | Pooled<br>(N = 11,642) | Vegetarians<br>(N = 2,658) | Non-vegetarians<br>(N = 8,984) |
| --- | --- | --- | --- |
| Age, mean (SD) | 48.5 (20.0) | 48.7 (20.6) | 48.4 (19.8) |
| Sex, N (%) |  |  |  |
| *Female* | 5,967 (51.2%) | 1,405 (52.9%) | 4,562 (50.8%) |
| *Male* | 5,675 (48.8%) | 1,253 (47.1%) | 4,422 (49.2%) |
| Years of schooling, mean (SD) | 7.0 (4.4) | 5.3 (4.0) | 7.5 (4.3) |
| BMI, mean (SD) | 22.5 (4.0) | 22.4 (3.9) | 22.5 (4.1) |
| Urban, N (%) | 3,910 (33.6%) | 406 (15.3%) | 3,504 (39.0%) |
| Rural, N (%) | 7,732 (66.4%) | 2,252 (84.7%) | 5,480 (61.0%) |
| Province, N (%) |  |  |  |
| *Liaoning* | 1,244 (10.6%) | 284 (10.6%) | 960 (10.6%) |
| *Heilongjiang* | 1,245 (10.6%) | 511 (19.2%) | 734 (8.2%) |
| *Jiangsu* | 1,264 (10.8%) | 116 (4.4%) | 1,148 (12.7%) |
| *Shandong* | 1,155 (9.9%) | 218 (8.2%) | 937 (10.4%) |
| *Henan* | 1,404 (12.0%) | 657 (24.7%) | 747 (8.3%) |
| *Hubei* | 1,249 (10.7%) | 348 (13.0%) | 901 (10.0%) |
| *Hunan* | 1,208 (10.3%) | 41 (1.5%) | 1,167 (12.9%) |
| *Guangxi* | 1,475 (12.6%) | 30 (1.1%) | 1,445 (16.0%) |
| *Guizhou* | 1,398 (12.0%) | 453 (17.0%) | 945 (10.5%) |

#### Cross-validation

To validate the phenotypes associated with vegetarianism identified through PheWAS, we conducted both logistic regression (for binary outcomes) and multivariate linear regression (for continuous outcomes) across all 443 phenotypes, comprising 9,009 vegetarian cases and 486,962 non-vegetarian controls. These regression models adjusted for key covariates, including sex, age, and the first ten genetic principal components, to control for potential confounding factors such as population stratification. We employed a Bonferroni correction to account for multiple comparisons across the 443 associations, maintaining a significance threshold of α = 0.05 to minimize Type I errors.

#### Mendelian Randomization

We employed a univariate Mendelian randomization approach using individual-level data to assess potential causal relationships between vegetarianism and various outcomes. Mendelian randomization’s strength lies in the random allocation of genes during meiosis, analogous to random treatment assignments in controlled trials, which may be impractical or unethical in this context. Only health and behavior-related traits identified as significant in both the PheWAS and cross-validation analyses (50 traits in total) were included in the MR analysis. Genetic instruments for the exposure were selected from the UKB cohort of 334,779 participants of European ancestry. Instruments were required to meet the following criteria: 1) genome-wide significance (*P* < 5×10^-6^); 2) absence of linkage disequilibrium (R^2^ < 0.01, window size = 10,000 kilobases [kb]); 3) exclusion of potential pleiotropic effects. *Post hoc* Sidak-Holm corrections were applied to all MR regressions to account for multiple comparisons. The STROBE-MR checklist is provided in the Supplemental Materials. The formal estimation followed a two-stage least squares (2SLS) specification:

*First stage:*

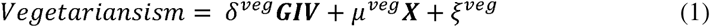

*Second stage:*

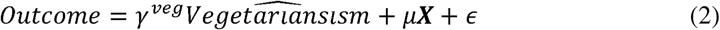

In addition, we conducted several sensitivity analyses to verify the causal relationships identified in one-sample MR analyses. First, a two-sample Mendelian randomization (2SMR) was performed using summary-level data from genome-wide association studies (GWAS). The primary exposure in the 2SMR was vegetarianism (GWAS Catalog ID: GCST90428838), with outcome data sourced from the Medical Research Council Integrative Epidemiology Unit (IEU) OpenGWAS database (https://gwas.mrcieu.ac.uk/). Detailed results are provided in Supplementary Tables 5. Second, Cochran’s Q tests were used to evaluate heterogeneity; if heterogeneity was present (*P* < 0.05), a random-effects inverse-variance weighted (IVW-RE) method was employed to address potential bias. Third, we performed MR-Egger intercept tests to assess horizontal pleiotropy. Where pleiotropy was detected (*P* < 0.05), the MR-PRESSO test was used to remove pleiotropic single nucleotide polymorphisms (SNPs), and pleiotropy-corrected results were generated. Lastly, leave-one-out (LOO) analyses were conducted to evaluate the consistency of the combined effect by sequentially removing each SNP, ensuring no individual SNP disproportionately influenced the results.

#### Machine-learning algorithms

We employed machine-learning algorithms to build a classification model to predict whether an individual is vegetarian (class 1) or not (class 0). Initially, we used LASSO regressions to select features from a total of 401 potential predictors, including 309 behavioral and physiological factors investigated earlier and an additional set of 92 genetic factors (see Supplementary Table 6 for details). After 100 iterations of LASSO with resampled training sets, we identified 43 factors that were selected in more than 50% of replications (Supplementary Table 7). These features were then used to construct two-class classification models using the Extreme Gradient Boosting (XGBoost) algorithm.[20] The models were trained using tenfold cross-validation. We assessed the model performance by calculating the area under the receiver operating characteristic curve (AUC), and utilized the SHapley Additive exPlanations (SHAP) algorithm to estimate the relative importance of the predictors affecting XGBoost’s vegetarianism estimates.[21]

## 3. Results

### Identifying health and behavioral outcomes for vegetarianism in PheWAS

In the PheWAS analysis, we investigated a total of 443 health and behavioral traits spanning nine groups as reported in Table 1a and Supplementary Table 1. Fig. 2a shows the geographic distributions of the average polygenic score for vegetarianism at the local authority level in UKB.

**Fig. 2.**
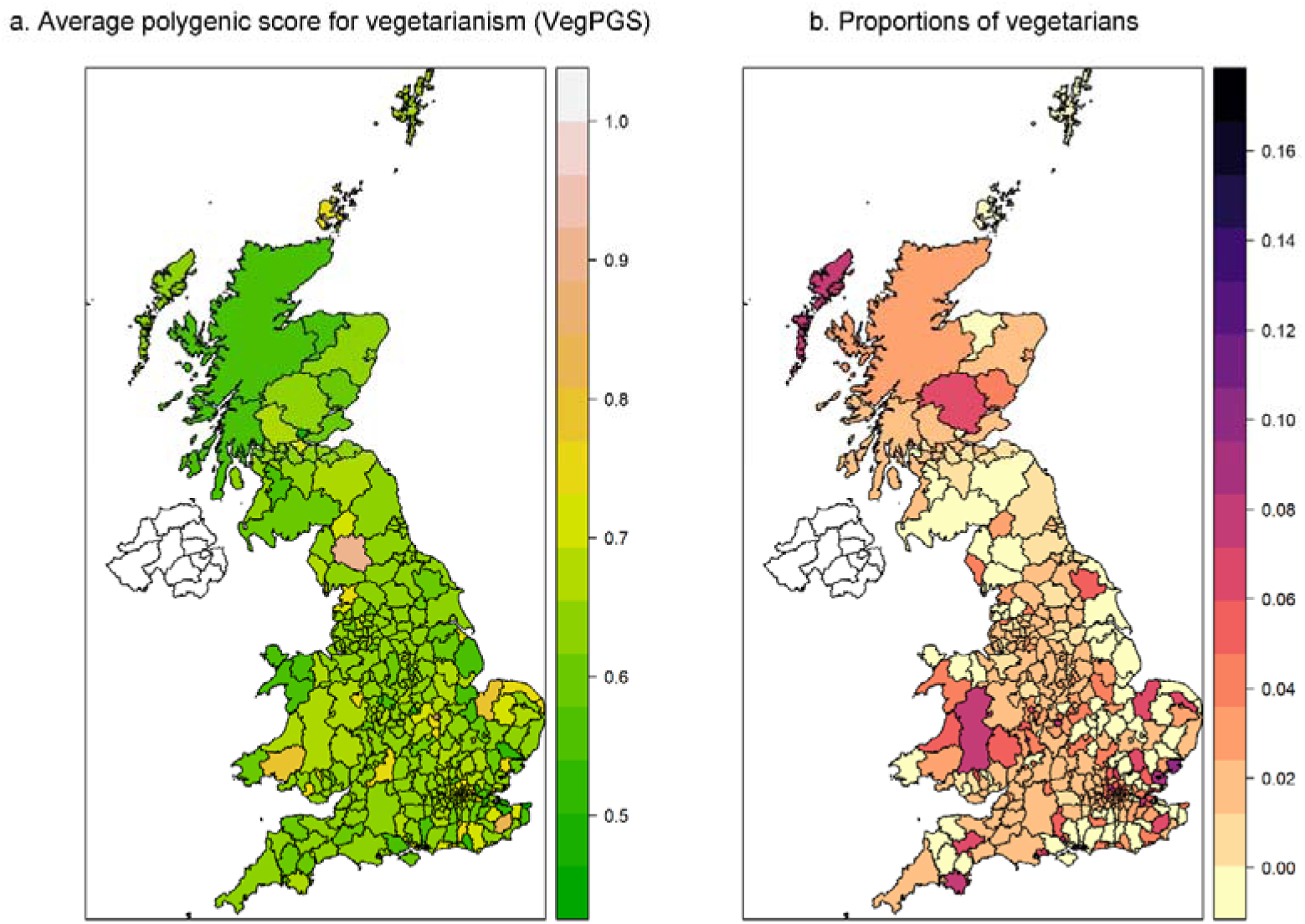
Geographic distributions of VegPGS and proportions of vegetarians in UKB.

In total, we found 57 health phenotypes and only 1 behavioral phenotype that showed significant associations with VegPGS after Bonferroni correction for multiple comparisons (*P* < 1.13×10^-4^; Fig. 3a and b). Detailed PheWAS results are presented in Supplementary Table 2.

**Fig. 3.**
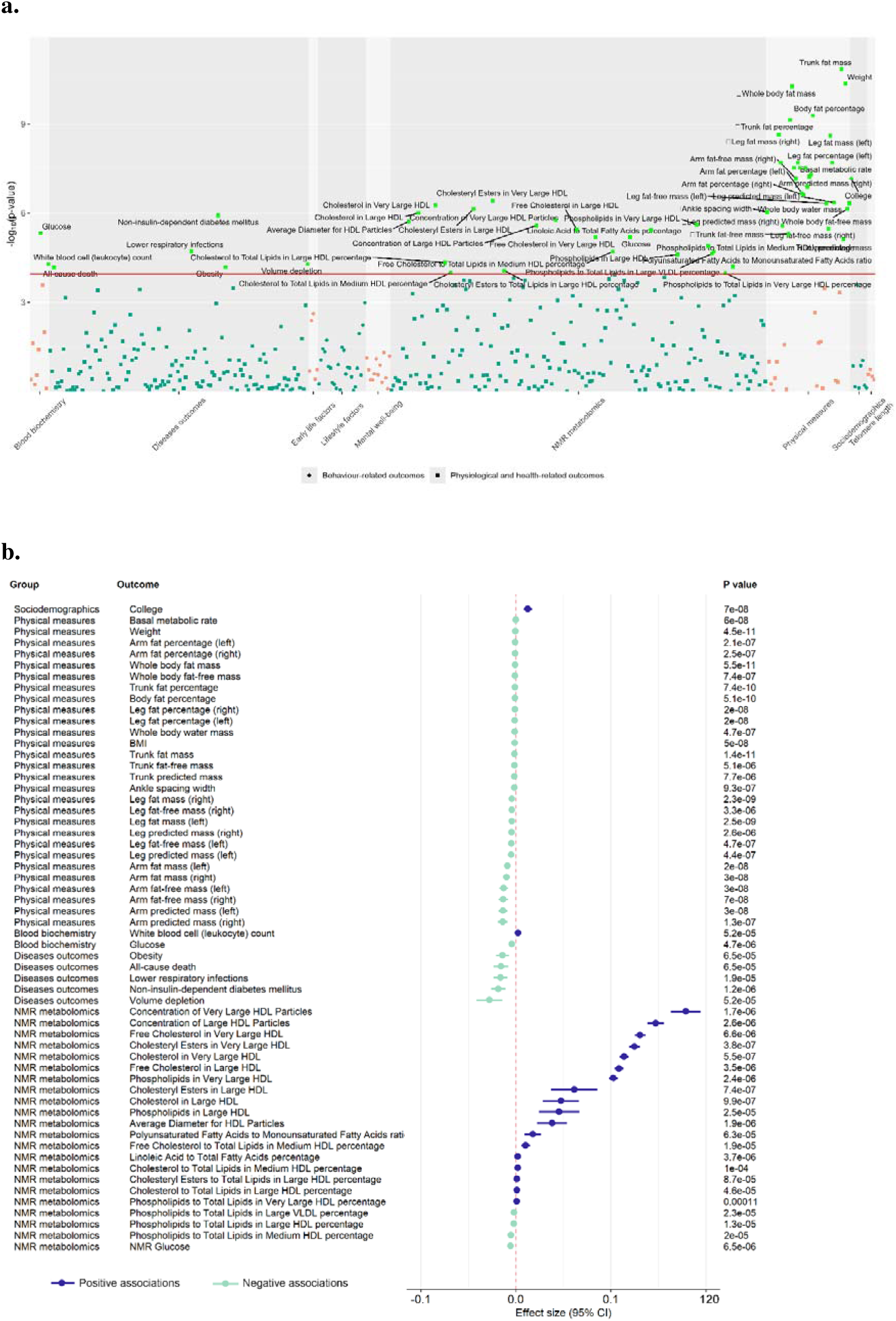
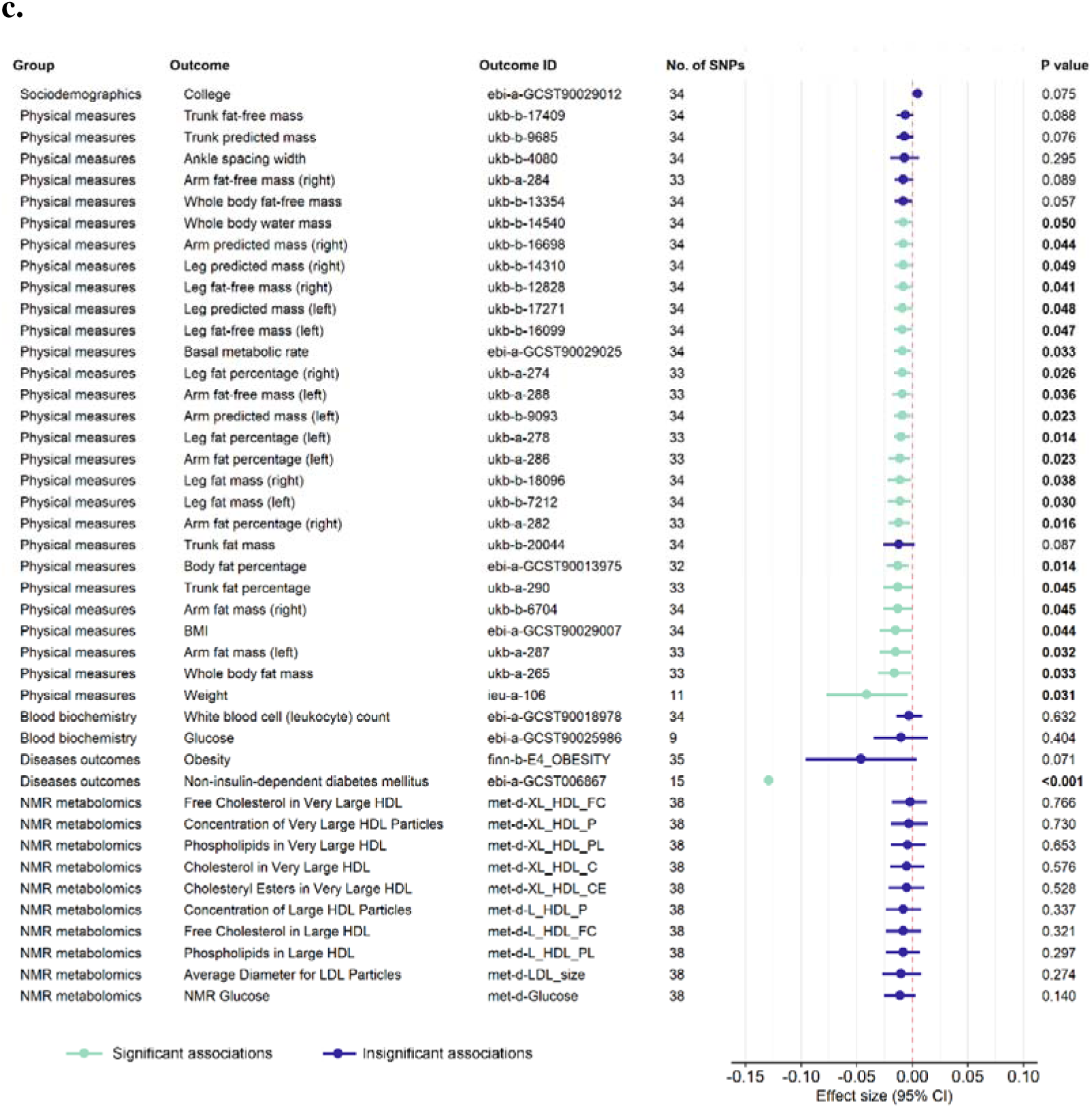
PheWAS and MR results. a. Manhattan plot of PheWAS. b. Forrest plot of significant associations after Bonferroni correction for multiple comparisons. c. Forrest plot of IVW-RE estimates from Mendelian randomization analyses.

Among behavioral phenotypes, higher VegPGS was only associated with a higher probability of having a college degree that remained significant after Bonferroni correction (*P* = 7.0×10^-8^). We did not find significant associations between VegPGS and early life factors, lifestyle factors, or mental well-being after corrections for multiple hypothesis testing.

In health-related phenotypes, VegPGS demonstrated Bonferroni-significant associations with 28 physical measures, 2 blood chemistry biomarkers, 5 disease outcomes, and 22 NMR metabolites. In specific, higher VegPGS was associated with lower basal metabolic rate (*P* = 6.0×10^-8^), which measures the minimum number of calories that an individual’s body needs to perform necessary functions. Besides, VegPGS showed an identical negative effect direction with 27 fat mass-related phenotypes, including weight, BMI, arm fat mass and percentages, leg fat mass and percentages, trunk fat mass and percentage, as well as whole body fat mass and fat-free mass (*P* = 1.45×10^-11^ to 7.71×10^-6^).

For disease outcomes, higher VegPGS were associated with lower risks of all-cause death (*P* = 6.52×10^-5^), obesity (*P* = 6.48×10^-5^), lower respiratory infections (*P* = 1.93×10^-5^), non-insulin-dependent diabetes mellitus (*P* = 1.19×10^-6^), and volume depletion (*P* = 5.23×10^-5^).

Regarding blood and metabolic biomarkers, VegPGS positively correlated with white blood cell count and several lipid-related NMR metabolites, such as concentration of large HDL particles, free cholesterol in large HDL, and phospholipids in very large HDL. Conversely, it was associated with lower glucose levels in both blood chemistry and NMR metabolomics.

### Cross-validation of related outcomes in case-control analysis

To validate the related health and behavioral outcomes identified in PheWAS, we performed multivariate linear and logistic regression analyses for all phenotypes on the UKB dataset containing 9,009 vegetarian cases and 486,962 non-vegetarian controls. Table 1b presents demographic characteristics of the vegetarian and non-vegetarian groups in UKB (54.4% female; mean [s.d.] age, 56.5 [8.1] years). Notably, 22.1% of vegetarian participants in UKB were from London, which is 9.6 percentage points higher than the overall regional average of 12.5%. Fig. 2b graphically shows the proportions of vegetarians at the local authority level in UKB.

In total, 49 health-related outcomes and 1 behavioral outcome remained significant in the case-control analysis after Bonferroni correction (*P* < 1.13×10^-4^). Supplementary Table 3 offers detailed results. Most associations identified in PheWAS were confirmed in the cross-validation analysis, demonstrating consistent effect directions. For example, vegetarianism was associated with a higher probability of having a college degree, reduced risks of obesity and non-insulin-dependent diabetes mellitus, lower basal metabolic rate, and lower fat mass-related phenotypes.

However, discrepancies were observed regarding white blood cell count and several lipid-related NMR metabolites (e.g., free cholesterol in large HDL), where case-control analysis indicated negative associations with vegetarianism, diverging from PheWAS findings. These inconsistencies underscore the complexity of the interactions between genetic predisposition and phenotypic expressions of vegetarian dietary patterns. Further investigation into these discrepancies is warranted to clarify the causality and underlying mechanisms.

### MR results on vegetarianism and health/behavioral outcomes

To further investigate the causal impacts of vegetarianism, we first conducted one-sample MR analyses on 50 phenotypes, significant in both PheWAS and cross-validation, using two-stage least squares. Out of these, 33 outcomes reached significance (*P* < 0.05), with 30 remaining significant after Sidak-Holm correction (Supplementary Table 4).

Next, we performed two-sample MR analyses for 43 phenotypes with available GWAS data. After accounting for heterogeneity and pleiotropy, 23 associations remained significant, confirming robust causal relationships (Fig. 3c). All robust associations pertained to health/physiological phenotypes. Detailed estimation results from the random-effects IVW method (robust to the presence of heterogeneity), heterogeneity tests with Cochran’s Q statistics, and MR-Egger intercept tests for horizontal pleiotropy were presented in Supplementary Tables 5. Scatter plots and LOO plots were presented in Supplementary Figures 1-43.

Notably, vegetarianism showed strong protective effects on non-insulin-dependent diabetes mellitus (beta = -0.1292, OR = 0.88, *P* = 5.70×10^-6^), lower BMI (beta = -0.0146, *P* = 0.0439), and lower basal metabolic rate (beta = -0.0088, *P* = 0.0327), along with reduced fat mass across 20 measures. However, no causal connections were found with educational attainment or obesity, challenging earlier observational findings. Additionally, no significant causal effects were noted on blood glucose or lipid-related NMR metabolites, suggesting prior inconsistency associations were likely spurious due to endogeneity bias.[22] This comprehensive approach, integrating one-sample and two-sample MR analyses, alongside rigorous sensitivity tests, strengthens the robustness of the findings while acknowledging the complexity of health outcomes linked to vegetarianism.

### Vegetarianism and health-related outcomes in East Asians

To evaluate the generalizability of our main findings, we analyzed the associations of vegetarianism with diabetes and BMI using available data from the CHNS cohort, which includes 2,658 vegetarians and 8,984 non-vegetarians. Table 1c summarizes demographic characteristics (51.2% female; mean age: 48.5 [20.0] years). Unlike the UKB cohort, CHNS vegetarians were less educated than their non-vegetarian counterparts (average years of schooling: 5.3 vs. 7.5). Regression analyses revealed significant negative associations of vegetarianism with diabetes (beta = -0.7766, OR = 0.46, *P* = 0.001) and BMI (beta = -0.2353, *P* = 0.020). These consistent findings provide triangulated evidence supporting our main results and enhance their applicability across different ethnic groups, reinforcing the potential global health benefits of vegetarianism.

### Machine-learning classification models for vegetarianism

To differentiate vegetarians from non-vegetarians, we developed machine-learning classification models using behavioral, physiological, and genetic factors. We initially considered 401 features (Supplementary Table 6), and through 100 replications of LASSO regression with resampled training set, we selected 43 key predictors that were included in more than 50% of the models (Supplementary Table 7). These included 11 behavioral factors (e.g., college education, current drinking status, family relationship satisfaction), 27 physiological factors (e.g., creatinine levels, hand grip strength, red blood cell count), and 5 genetic factors (e.g., polygenic scores for height, educational attainment, rs7012814).

Three XGBoost classification models were constructed: 1) a baseline model using only behavioral factors, 2) a model incorporating behavioral and physiological factors, and 3) a full model that also included genetic data. In the training cohort, the area under the receiver operating characteristic (AUC) curve for the baseline model was 0.744 (95% CI: 0.730 to 0.758, Supplementary Fig. 44). Adding physiological factors improved the AUC to 0.928 (95% CI: 0.921 to 0.935, Supplementary Fig. 45), and the full model incorporating genetic factors achieved an AUC of 0.932 (95% CI: 0.925 to 0.939, Fig. 4a).

**Fig. 4.**
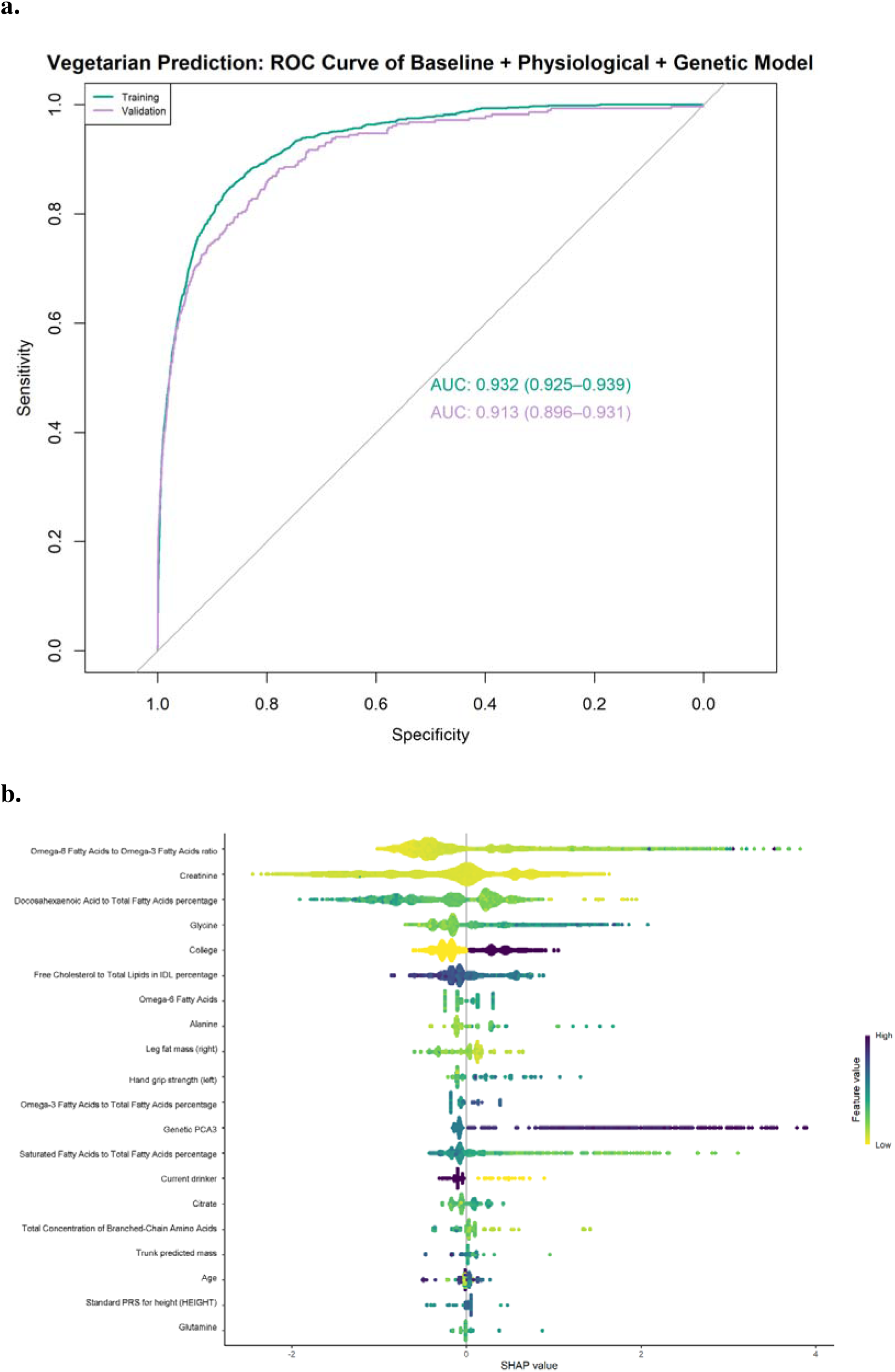
Performance and SHAP visualization of machine-learning models for vegetarian classification task. **a.** AUC plots for the ROC curves of the full classification model. **b.** SHAP beeswarm plot of top 20 predictors. Each participant is represented as a data point. The width of the horizontal bars reflects their impact on model predictions, with a wider range indicating a larger impact. The color of the horizontal bars represents the magnitude of predictors, which was coded in a gradient from yellow (low) to dark purple (high), shown as the color bar on the right-hand side. The x-axis indicates the likelihood of being a vegetarian (right) or non-vegetarian (left). Using the predictor ‘college’ as an example, those with a college degree (colored dark purple) are more likely to be vegetarians (right side), while those without a college degree (colored yellow) tend to be non-vegetarians (left side).

In the validation cohort, the full model still exhibited strong predictive power, with an AUC of 0.913 (95% CI: 0.896 to 0.931, Fig. 4a), compared to 0.728 (95% CI: 0.699 to 0.756, Supplementary Fig. 44) for the baseline model and 0.910 (95% CI: 0.892 to 0.927, Supplementary Fig. 45) for the model with behavioral and physiological factors. The SHAP analysis further identified key predictors, with the top five being the omega-6 to omega-3 fatty acids ratio, creatinine levels, docosahexaenoic acid percentage, glycine levels, and college education (Fig. 4b).

Overall, the full classification model demonstrated strong discrimination performance,[23] particularly when integrating behavioral, physiological, and genetic data, indicating that these combined factors are crucial in predicting vegetarianism with high accuracy.

## 4. Discussion

The results of this study highlight the multifaceted health and behavioral impacts of vegetarianism, providing valuable insights into both the positive and nuanced consequences of adopting this dietary pattern. One of the key findings is the robust protective effect of vegetarianism against non-insulin-dependent diabetes mellitus, lower BMI, and reduced fat mass across multiple body regions. These results corroborate prior observational studies that have highlighted the health benefits of plant-based diets in reducing metabolic risks. The causal evidence provided by MR analyses and replications in East Asian populations further strengthens these associations, moving beyond mere correlation to suggest that vegetarianism actively contributes to better metabolic health outcomes across different ethnic groups. In the exploration of using machine-learning models to predict vegetarianism, the inclusion of physiological and genetic factors significantly improved model accuracy, implying that vegetarianism may not merely a lifestyle choice, but one influenced by complex biological mechanisms.

Our findings on the protective effect of vegetarianism against type 2 diabetes align with prior research,[24–25] which consistently links plant-based diets to reduced diabetes risk due to their high fiber and low saturated fat content. This study adds causal and cross-population evidence to these observations, reinforcing the benefits of vegetarianism for metabolic health.

The causal association with lower basal metabolic rate (BMR) is intriguing, suggesting that vegetarianism may contribute to energy adaptation by reducing caloric needs. While a lower BMR implies fewer calories are required for essential functions,[26] it also presents challenges for individuals transitioning to vegetarianism, potentially necessitating personalized nutritional guidance to avoid energy imbalances.

Notably, our results did not find causal links between vegetarianism and cancer or cardiovascular disease risks, challenging earlier studies that suggested protective effects against conditions such as colorectal and breast cancers.[4–5,7,27] This discrepancy may stem from the limitations of observational studies, where confounding variables were not adequately addressed, as highlighted by the inconsistencies in glucose and lipid metabolism results. The use of MR allowed us to mitigate these confounding effects, revealing that previous associations with vegetarianism and metabolic markers may have been biased due to endogeneity.

Similarly, while previous research has reported varying associations between vegetarianism and mental health[8,28]—some indicating increased risks of depression and anxiety—our study did not find significant causal links between vegetarianism and mental health outcomes. This discrepancy may reflect the complexity of dietary patterns’ psychological impacts, warranting further investigation.

Some limitations should be acknowledged. While our MR approach strengthens causal inference, the lack of significant associations with certain traits (e.g., cancer risk) suggests that the protective effects of vegetarianism may be more limited than previously assumed. Additionally, our models are based on UK Biobank data, and though we replicated key findings in an East Asian population (CHNS), further validation across diverse populations is necessary to enhance the generalizability of these results.

In conclusion, this study advances our understanding of the health and behavioral impacts of vegetarianism by integrating multimodal health, physiological, behavioral, and genetic data. Future research should continue to explore the biological underpinnings of dietary behavior while expanding the scope to include diverse populations and more nuanced health and behavioral outcomes.

### Declaration of generative AI and AI-assisted technologies in the writing process

During the preparation of this work the authors used ChatGPT in order to improve the readability and language of the manuscript. After using this tool/service, the authors reviewed and edited the content as needed and take full responsibility for the content of the publication.

## Data Availability

Data described in the manuscript will be made available upon application and approval from the UK Biobank. For this study, permission to access and analyse the UK Biobank data was approved under the application 89068.

## Notes

### Competing Interest Statement

The authors have declared no competing interest.

### Author Declarations

UK Biobank is the database for a population-based study involving more than 500,000 UK residents approved by the NHS National Research Ethics Service (Ref: 11/NW/0382). Ethical approval for CHNS was granted by the institutional review committees of the University of North Carolina at Chapel Hill and National Institute for Nutrition and Health, Chinese Center for Disease Control and Prevention.

