## Supplementary Figures for "Triangulating evidence from the UK Biobank and China shows the health and behavioral impact of vegetarianism"

**Supplemental Materials:**

**1. Supplementary Figures 1-45**

**2. STROBE-MR checklist**

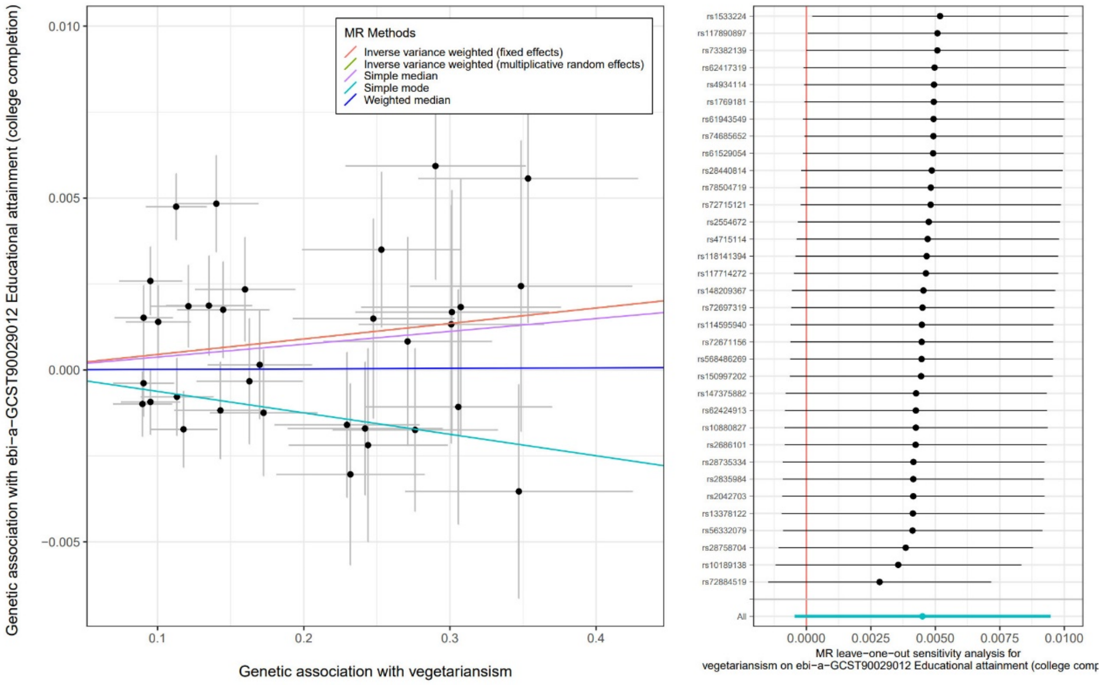


**Supplementary Fig. 1.** Scatter plot for the causal association between vegetarianism and educational attainment (college completion) and leave-one-out tests

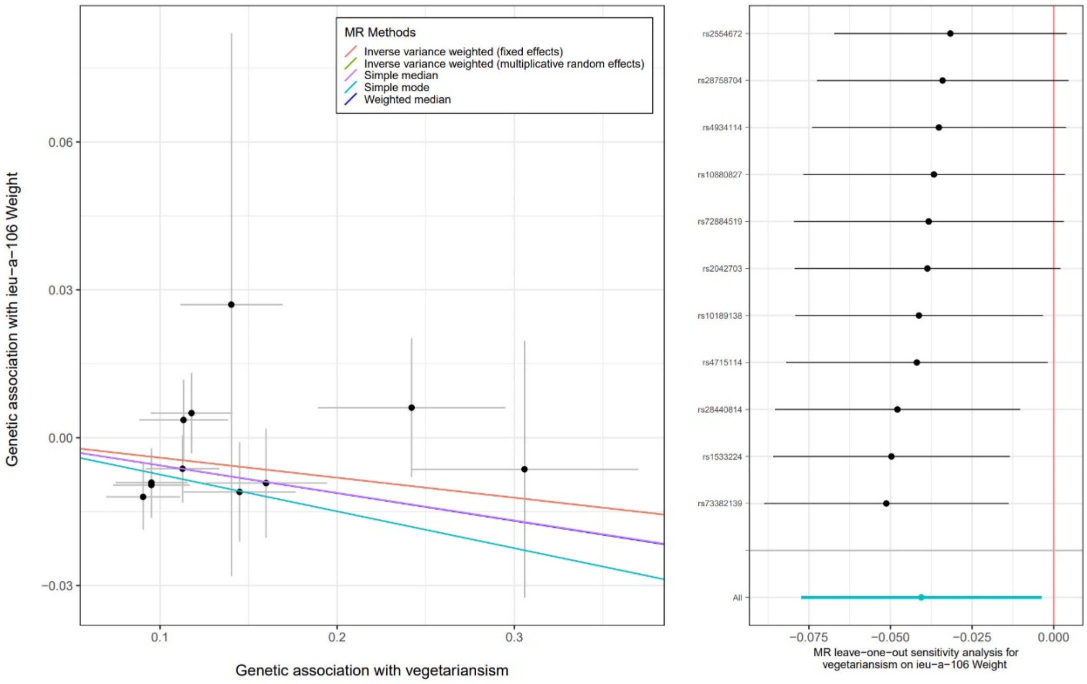


**Supplementary Fig. 2.** Scatter plot for the causal association between vegetarianism and weight and leave-one-out tests

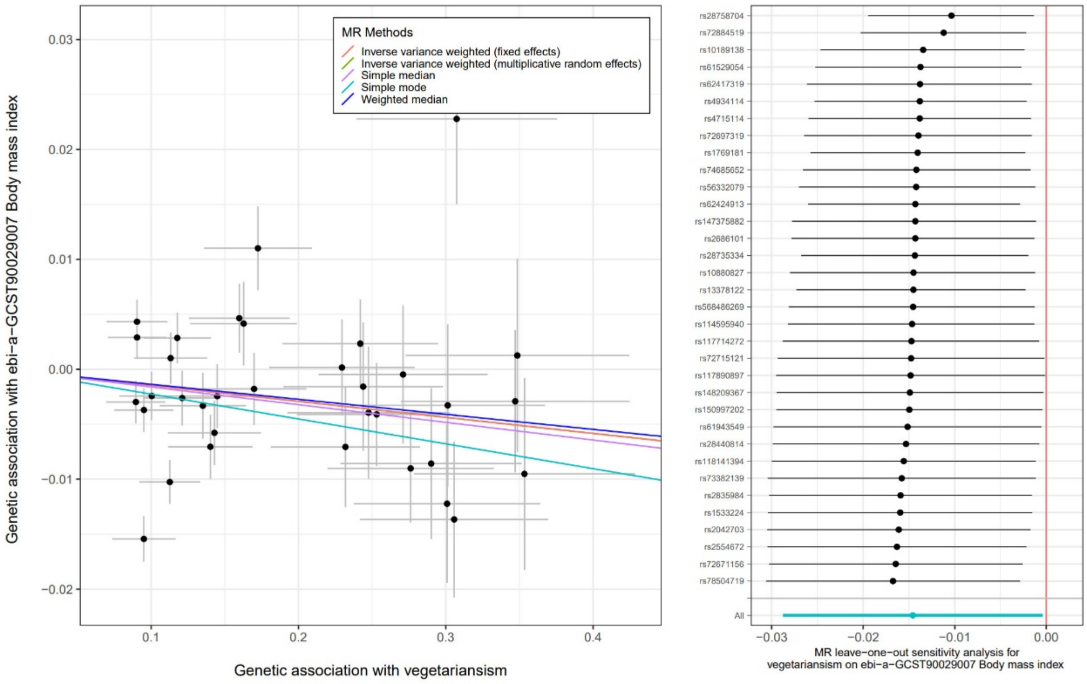


**Supplementary Fig. 3.** Scatter plot for the causal association between vegetarianism and body mass index and leave-one-out tests

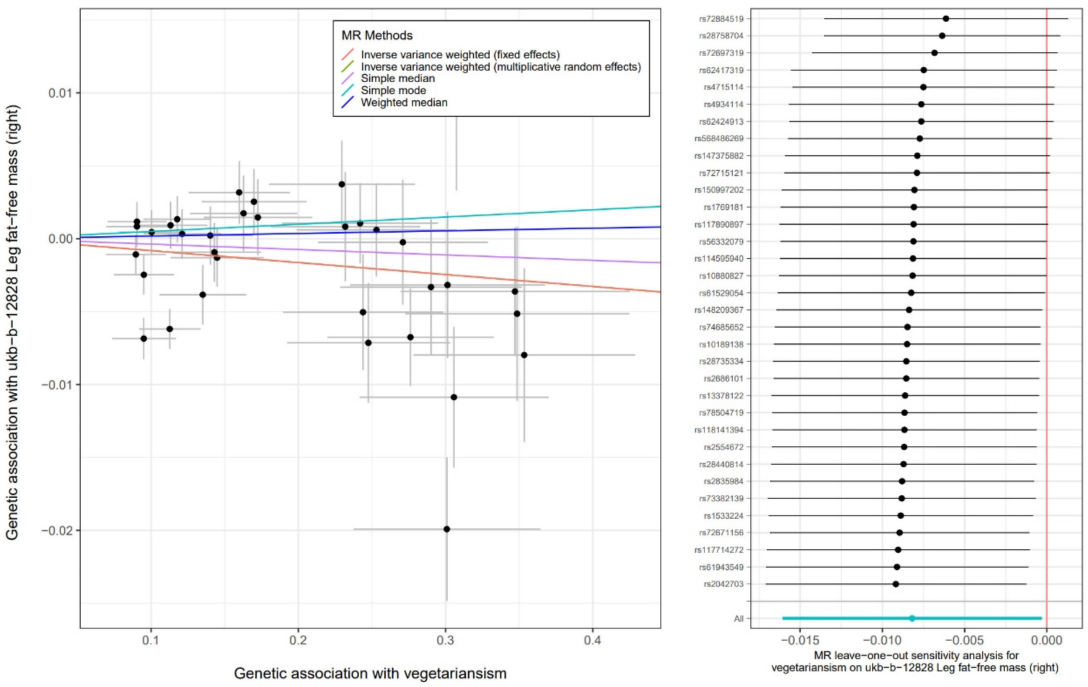


**Supplementary Fig. 4.** Scatter plot for the causal association between vegetarianism and leg fat-free mass (right) and leave-one-out tests

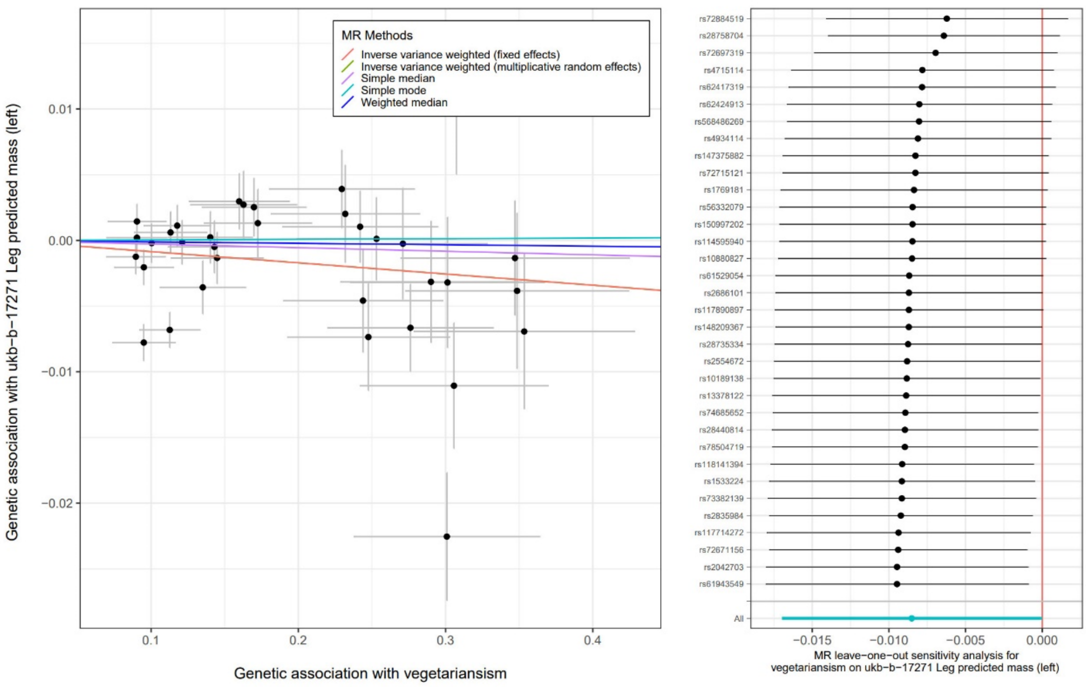

**Supplementary Fig. 5.** Scatter plot for the causal association between vegetarianism and leg predicted mass (left) and leave-one-out tests

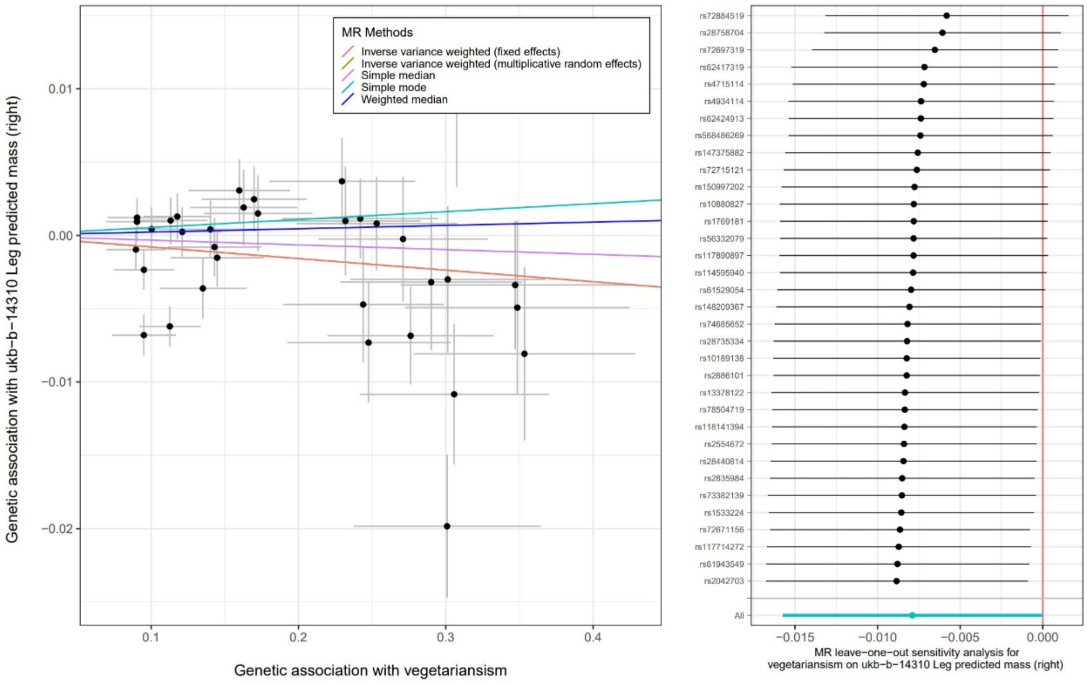


**Supplementary Fig. 6.** Scatter plot for the causal association between vegetarianism and leg predicted mass (right) and leave-one-out tests

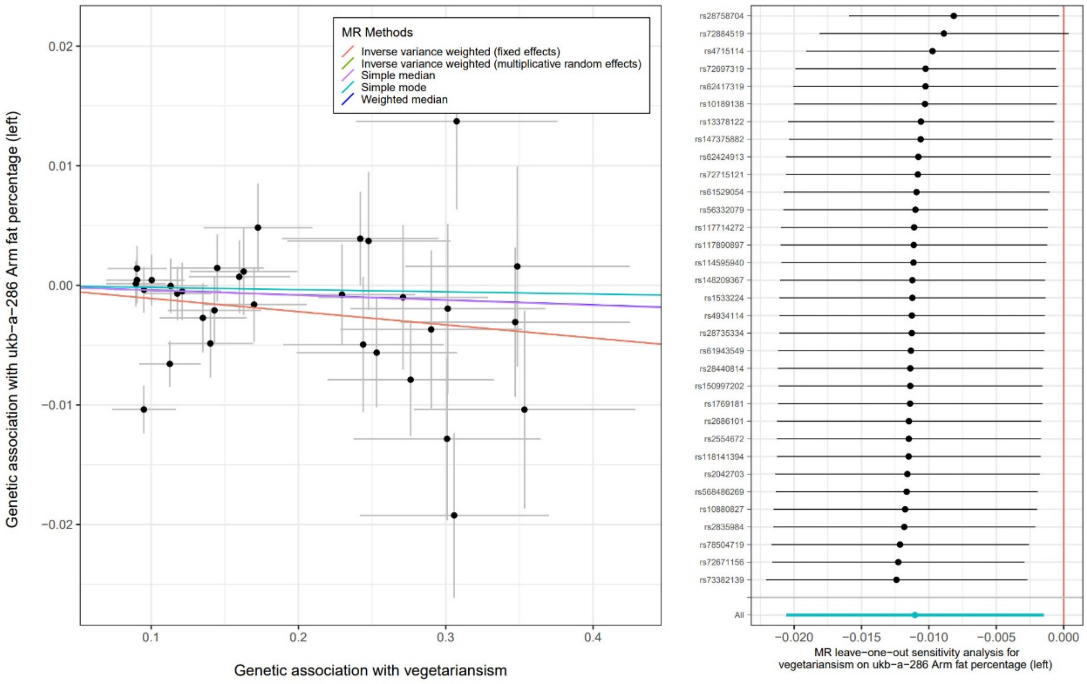


**Supplementary Fig. 7.** Scatter plot for the causal association between vegetarianism and arm fat percentage (left) and leave-one-out tests

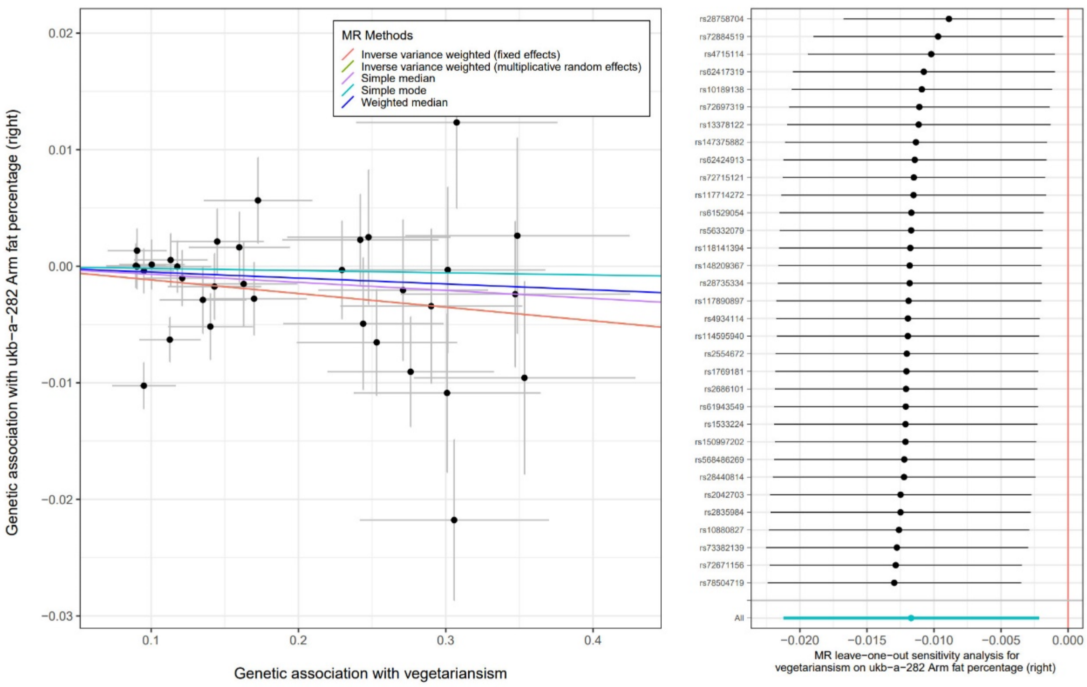


**Supplementary Fig. 8.** Scatter plot for the causal association between vegetarianism and arm fat percentage (right) and leave-one-out tests

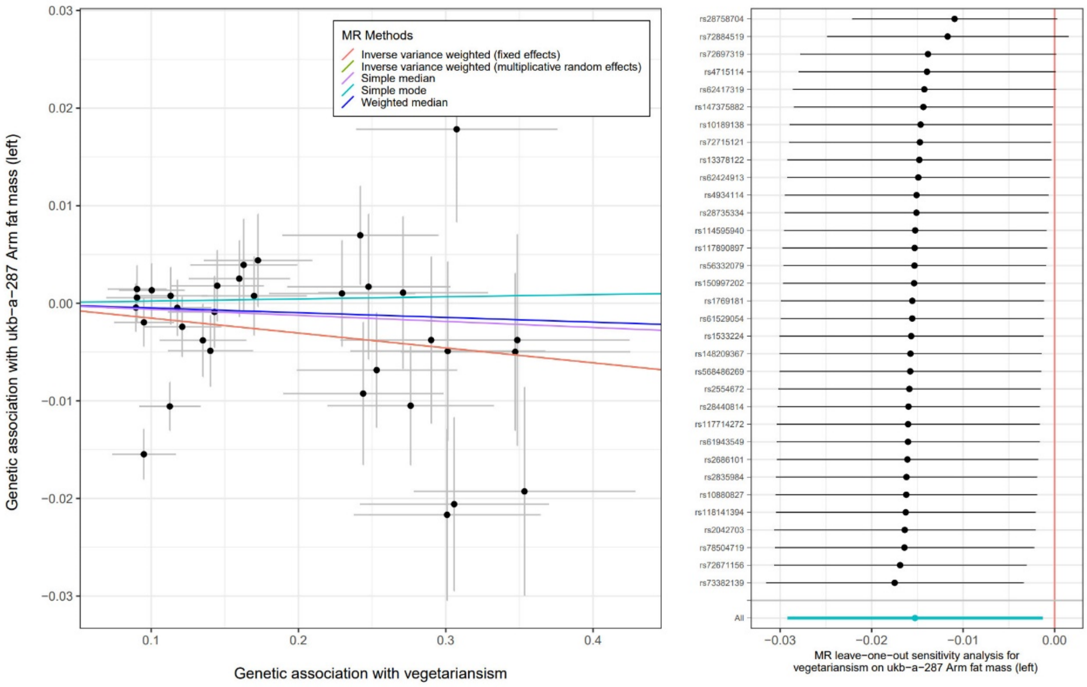


**Supplementary Fig. 9.** Scatter plot for the causal association between vegetarianism and arm fat mass (left) and leave-one-out tests

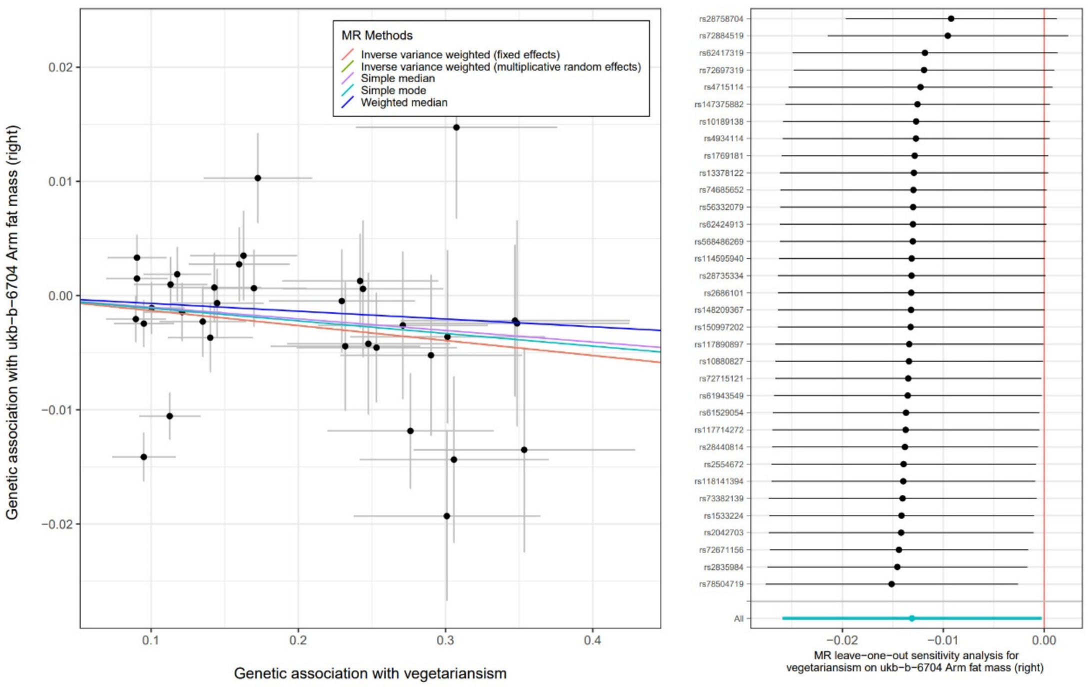


**Supplementary Fig. 10.** Scatter plot for the causal association between vegetarianism and arm fat mass (right) and leave-one-out tests

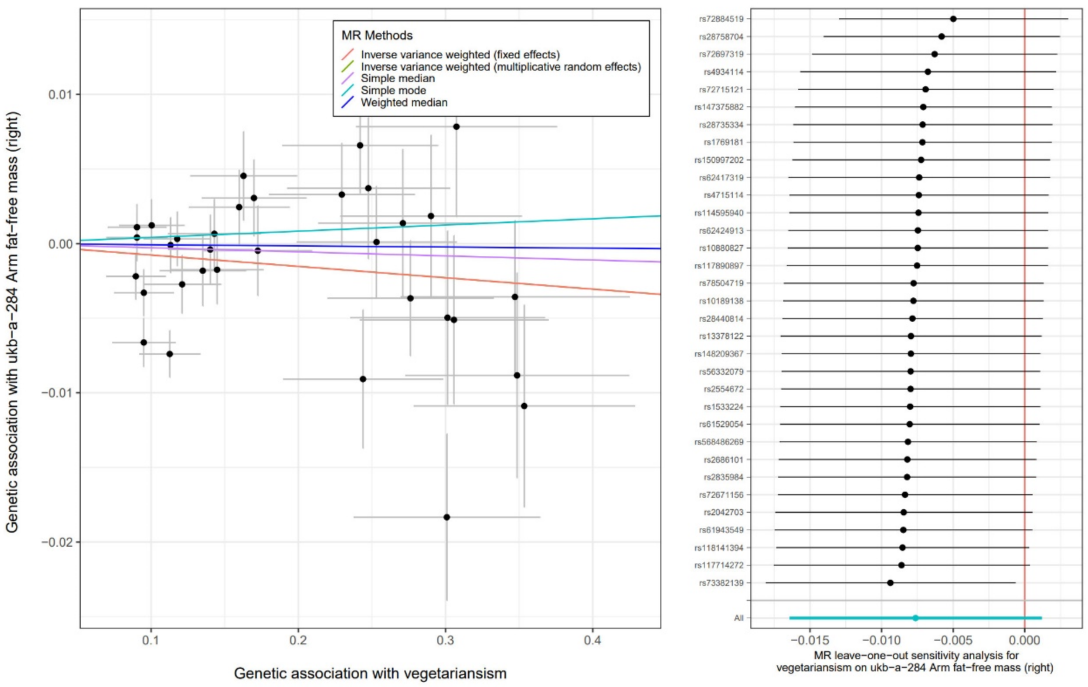


**Supplementary Fig. 11.** Scatter plot for the causal association between vegetarianism and arm fat-free mass (right) and leave-one-out tests

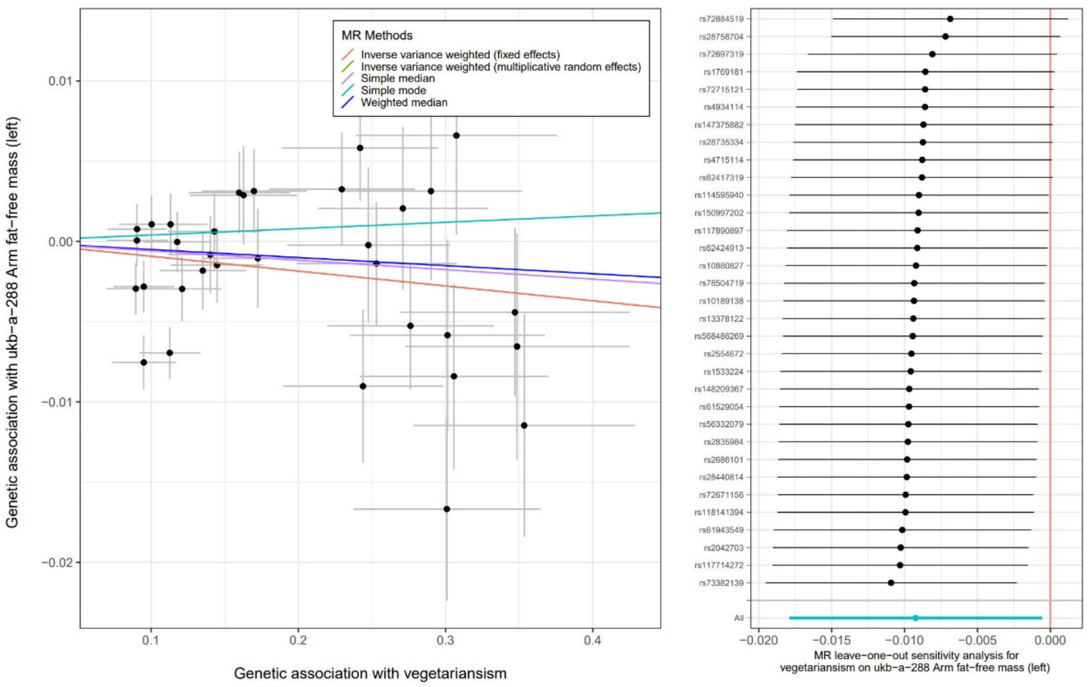


**Supplementary Fig. 12.** Scatter plot for the causal association between vegetarianism and arm fat-free mass (left) and leave-one-out tests

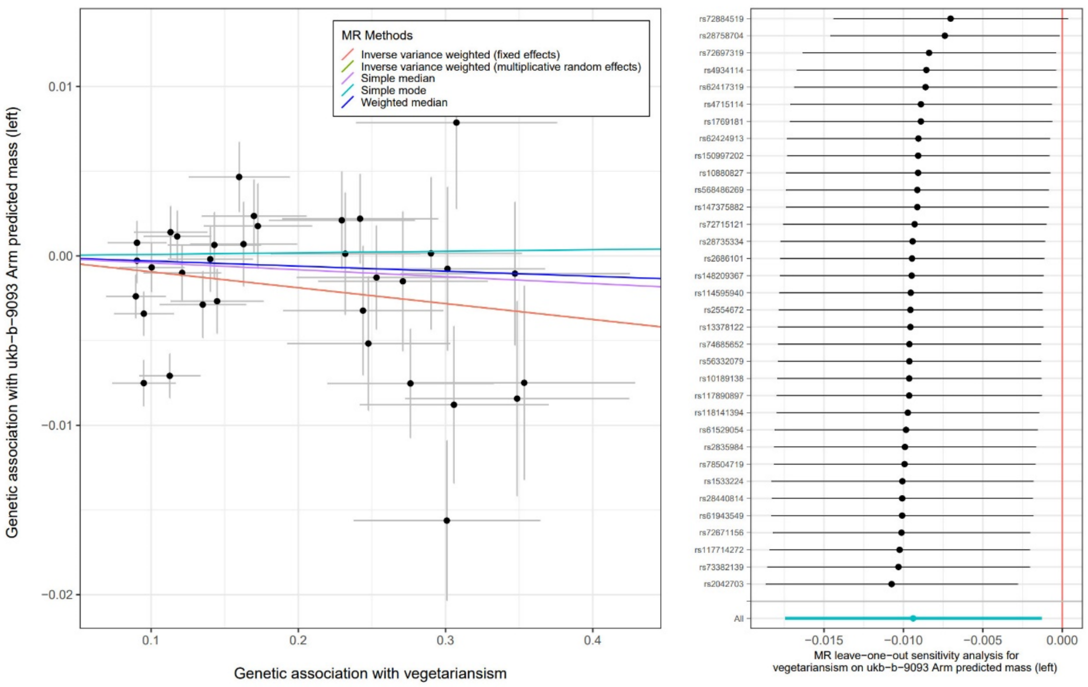


**Supplementary Fig. 13.** Scatter plot for the causal association between vegetarianism and arm predicted mass (left) and leave-one-out tests

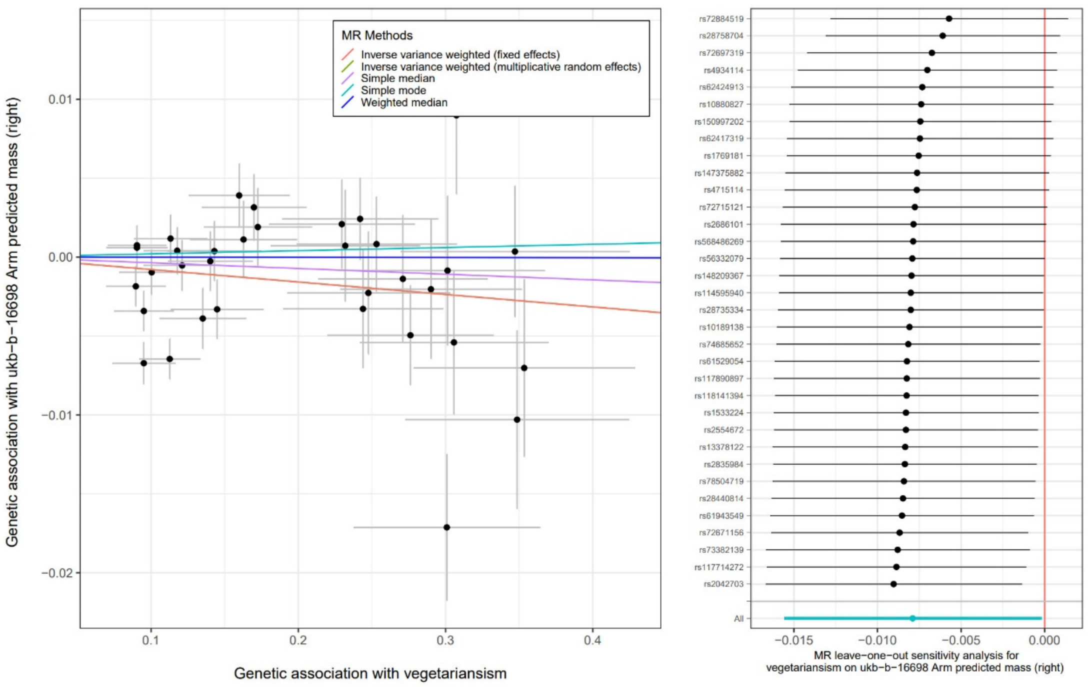


**Supplementary Fig. 14.** Scatter plot for the causal association between vegetarianism and arm predicted mass (right) and leave-one-out tests

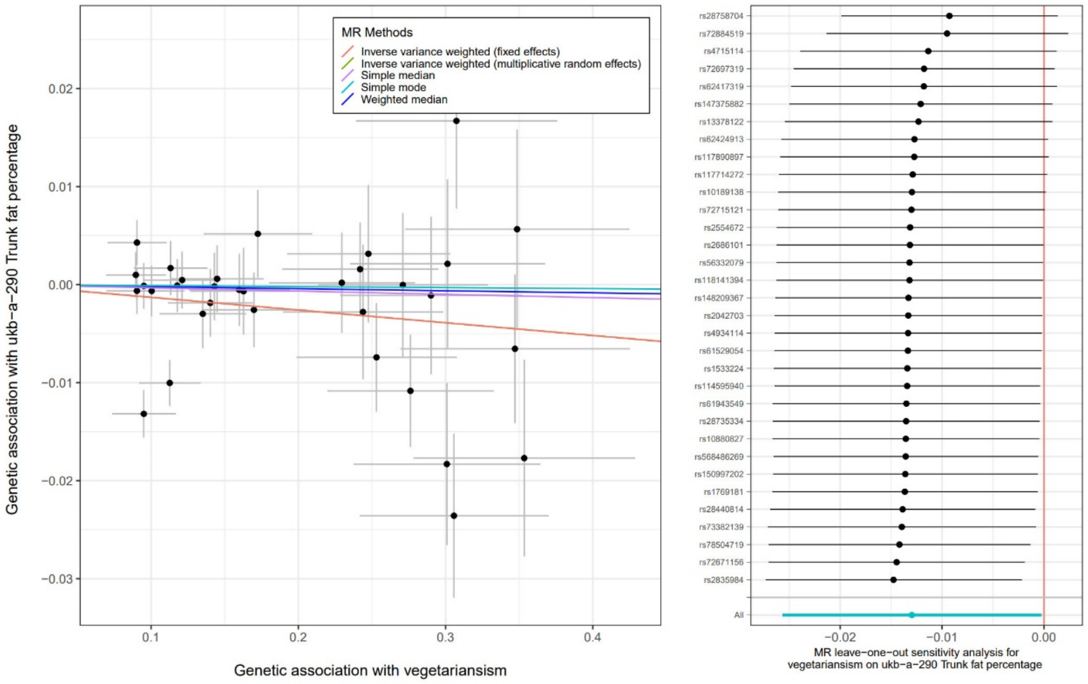


**Supplementary Fig. 15.** Scatter plot for the causal association between vegetarianism and trunk fat percentage and leave-one-out tests

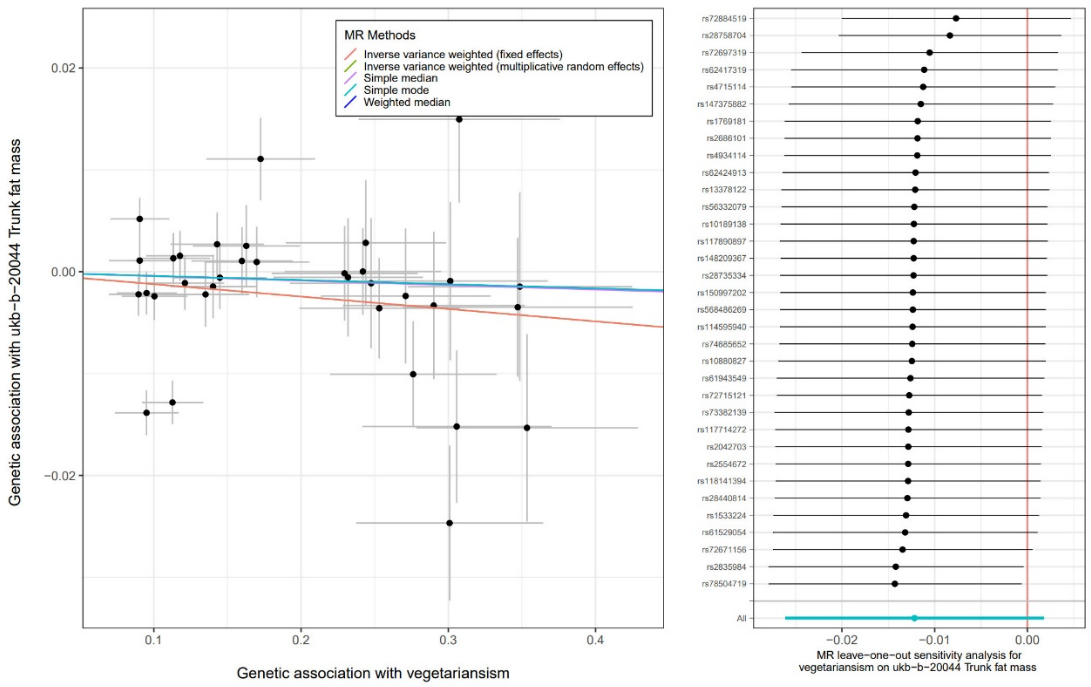


**Supplementary Fig. 16.** Scatter plot for the causal association between vegetarianism and trunk fat mass and leave-one-out tests

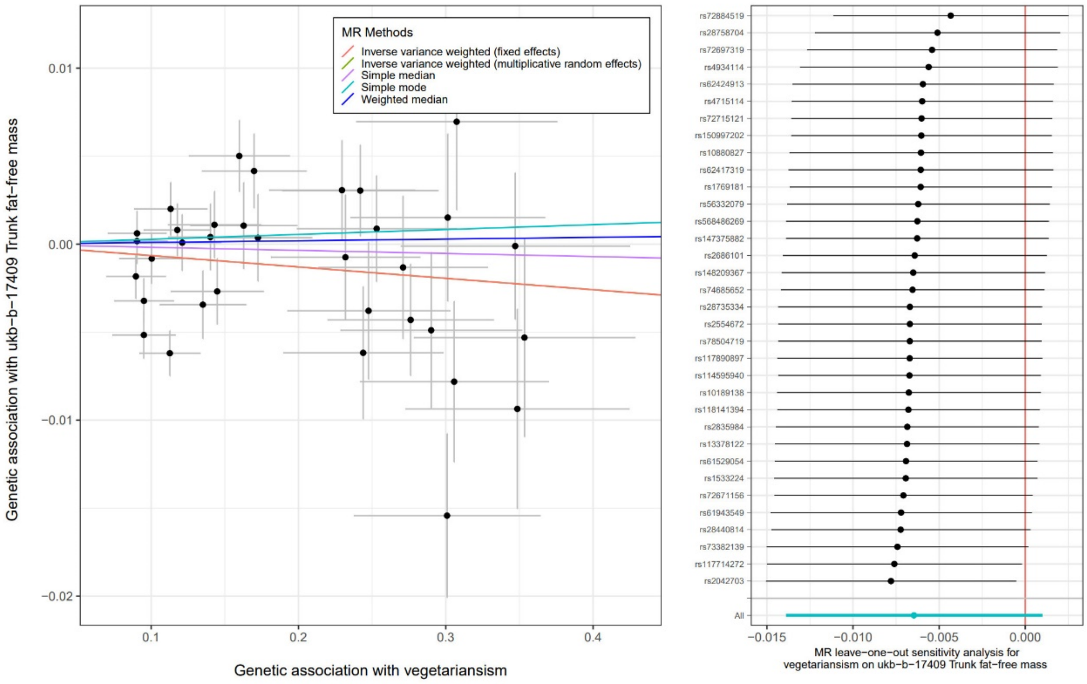


**Supplementary Fig. 17.** Scatter plot for the causal association between vegetarianism and trunk fat-free mass and leave-one-out tests

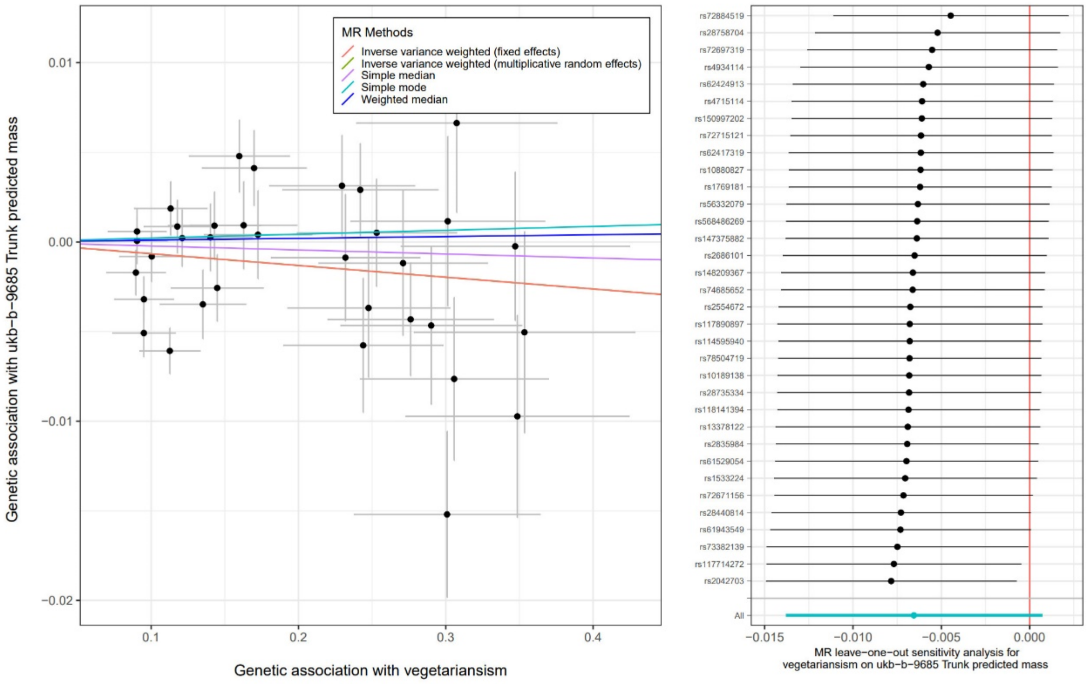


**Supplementary Fig. 18.** Scatter plot for the causal association between vegetarianism and trunk predicted mass and leave-one-out tests

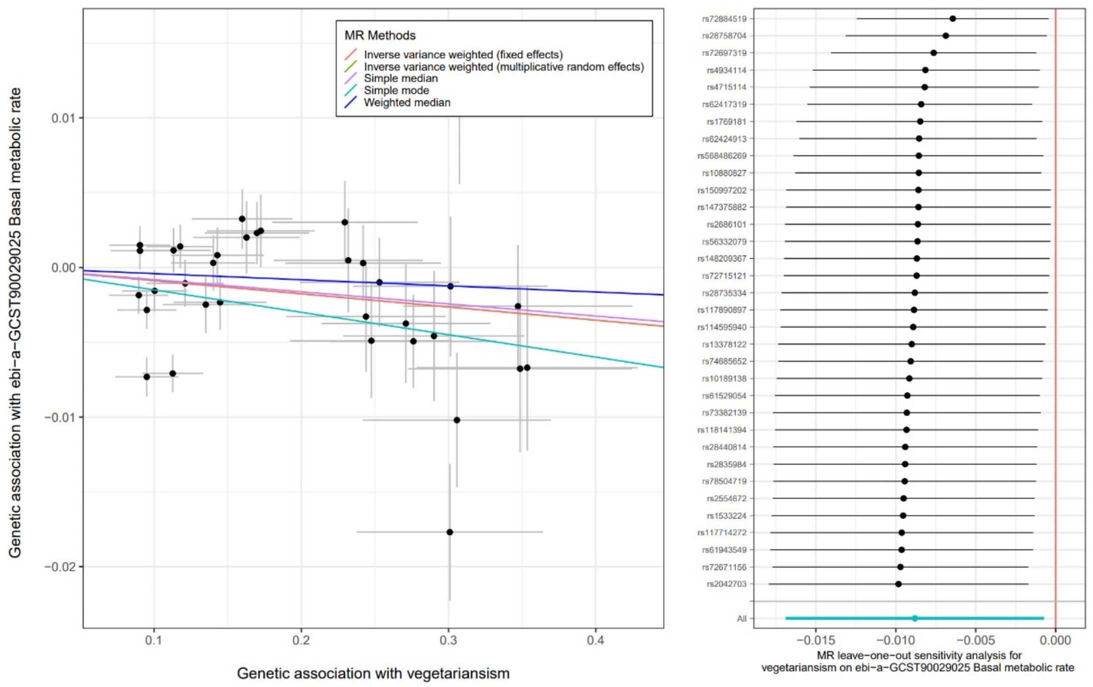


**Supplementary Fig. 19.** Scatter plot for the causal association between vegetarianism and basal metabolic rate and leave-one-out tests

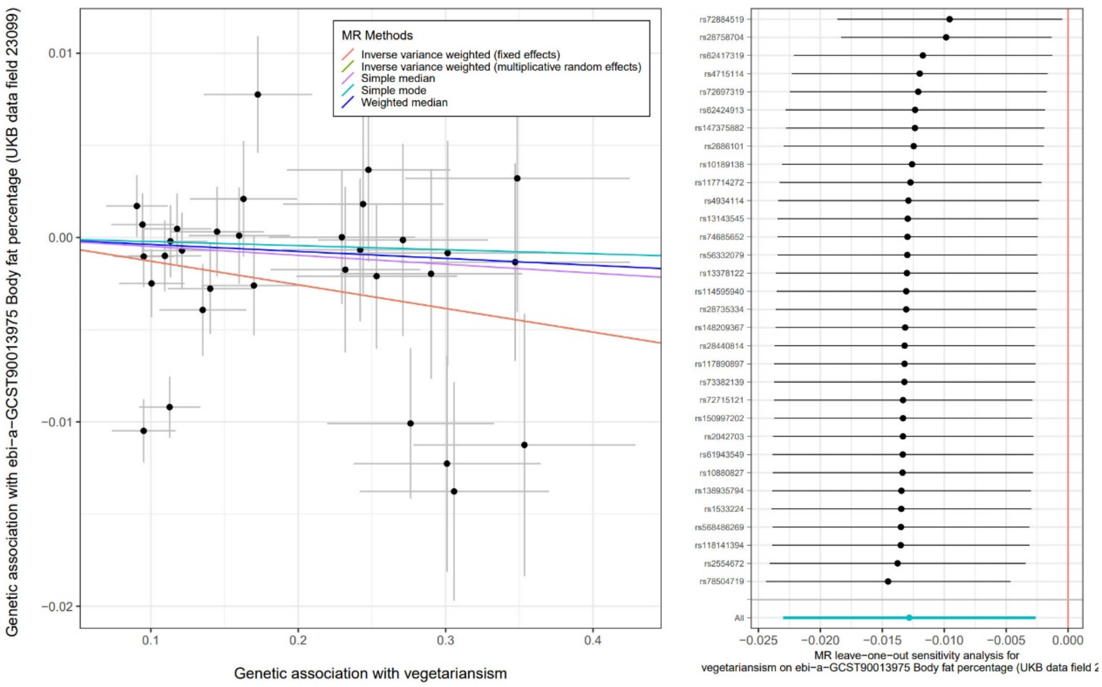


**Supplementary Fig. 20.** Scatter plot for the causal association between vegetarianism and body fat percentage and leave-one-out tests

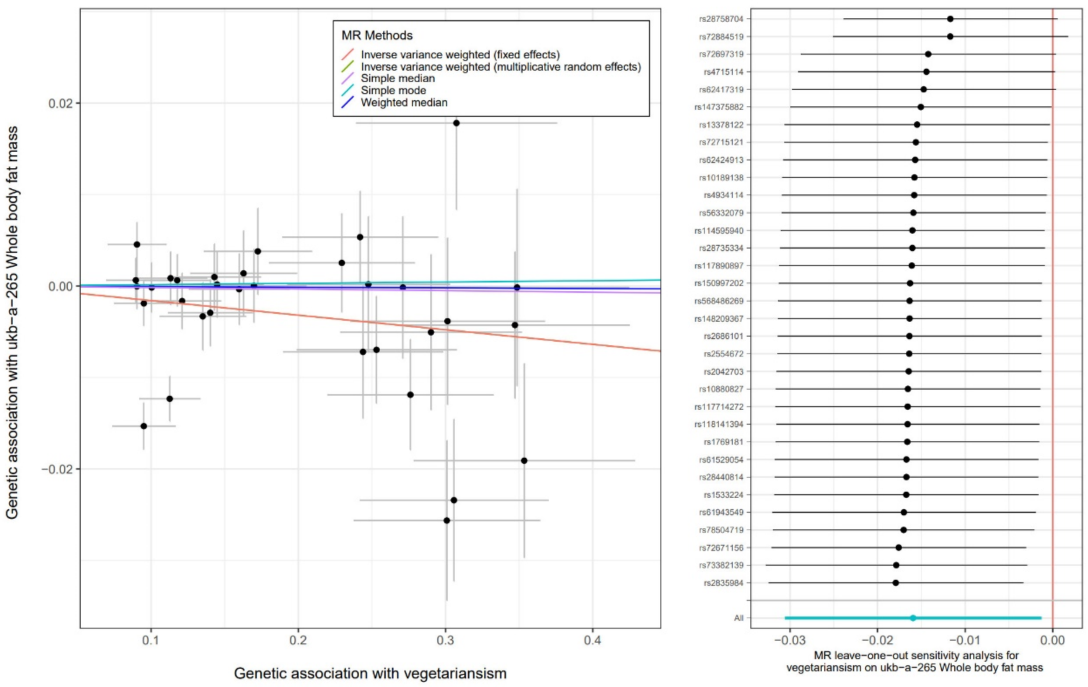


**Supplementary Fig. 21.** Scatter plot for the causal association between vegetarianism and whole body fat mass and leave-one-out tests

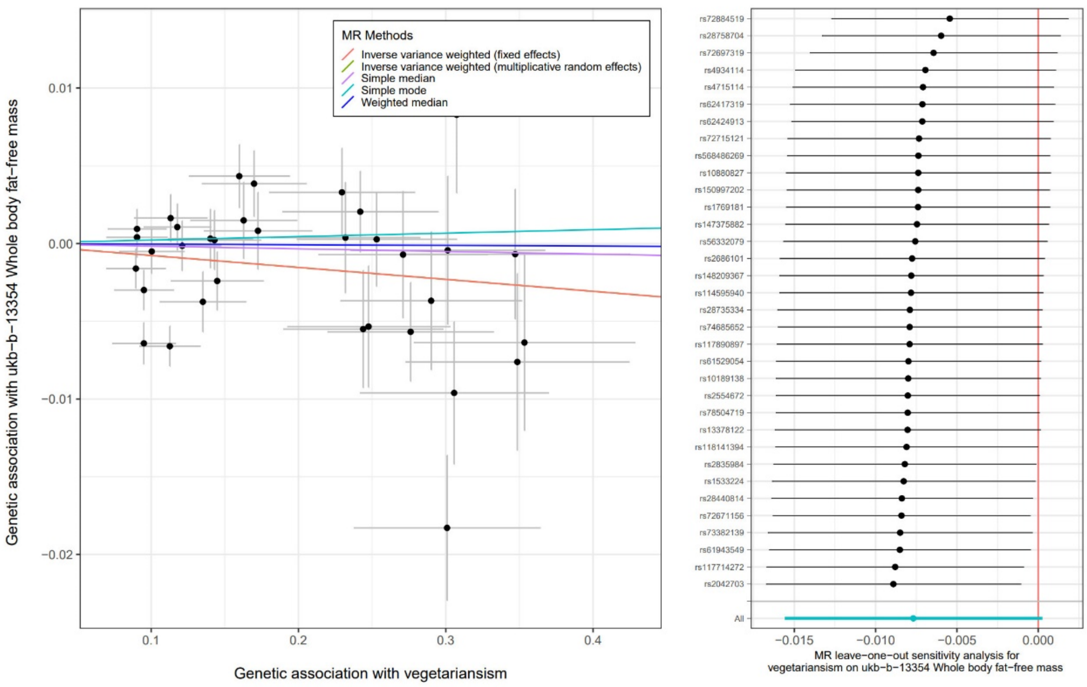


**Supplementary Fig. 22.** Scatter plot for the causal association between vegetarianism and whole body fat-free mass and leave-one-out tests

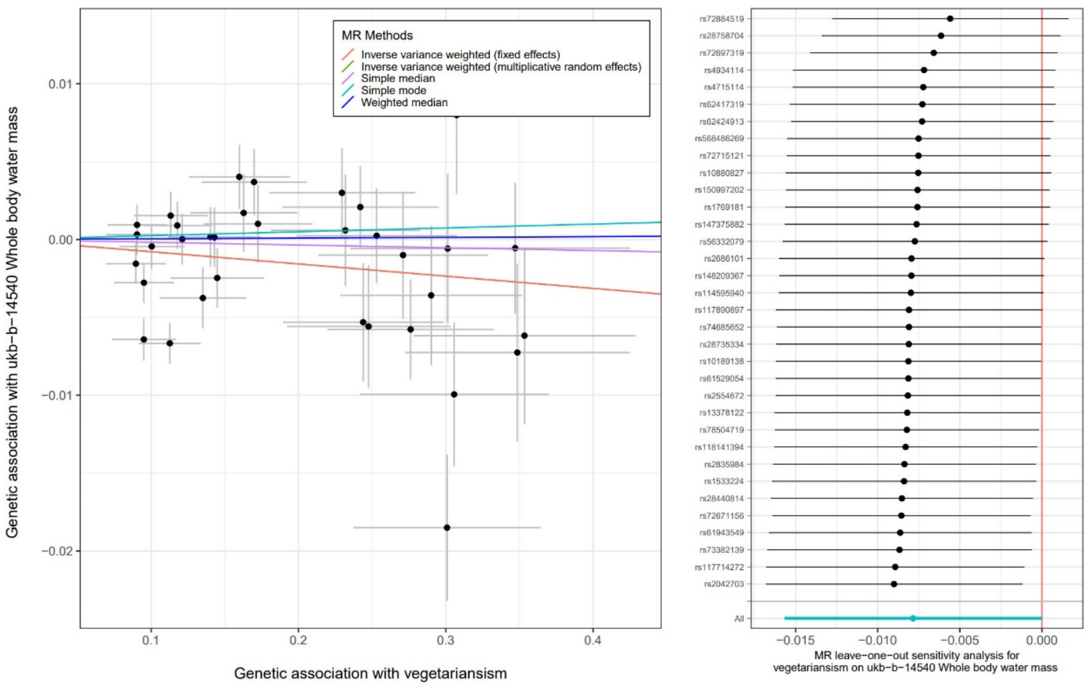


**Supplementary Fig. 23.** Scatter plot for the causal association between vegetarianism and whole body water mass and leave-one-out tests

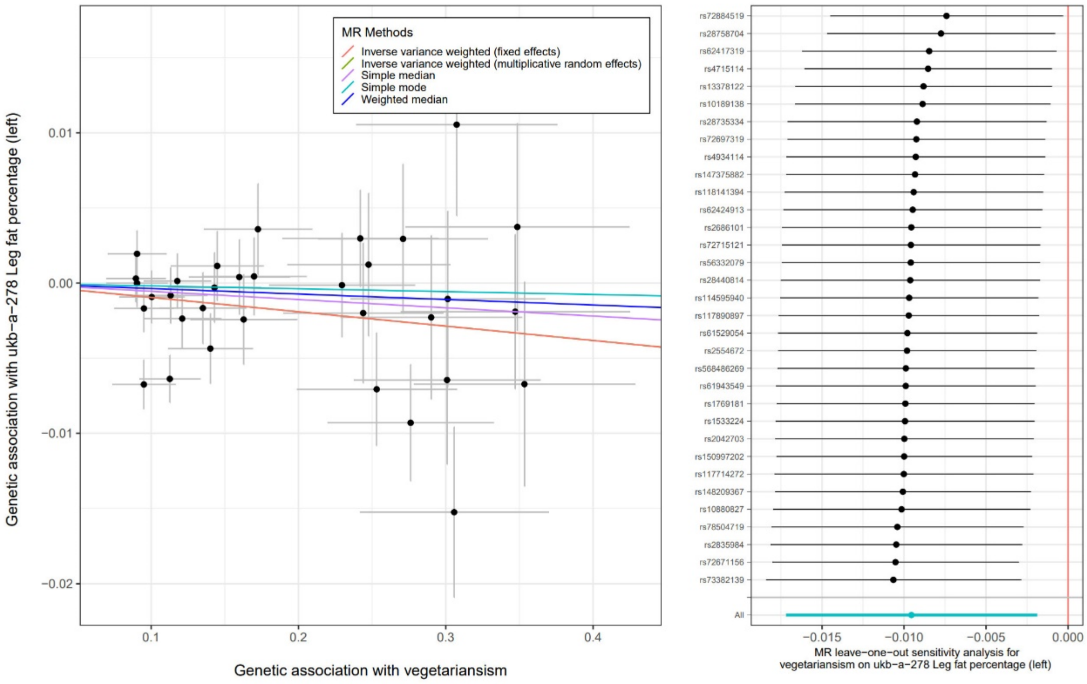


**Supplementary Fig. 24.** Scatter plot for the causal association between vegetarianism and leg fat percentage (left) and leave-one-out tests

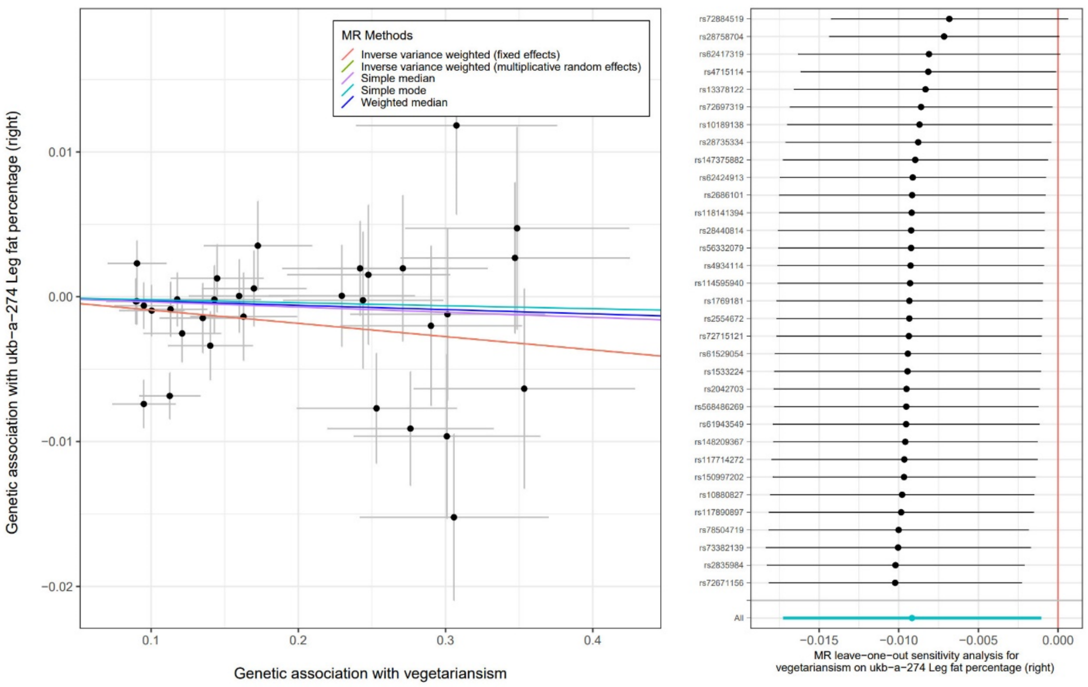


**Supplementary Fig. 25.** Scatter plot for the causal association between vegetarianism and leg fat percentage (right) and leave-one-out tests

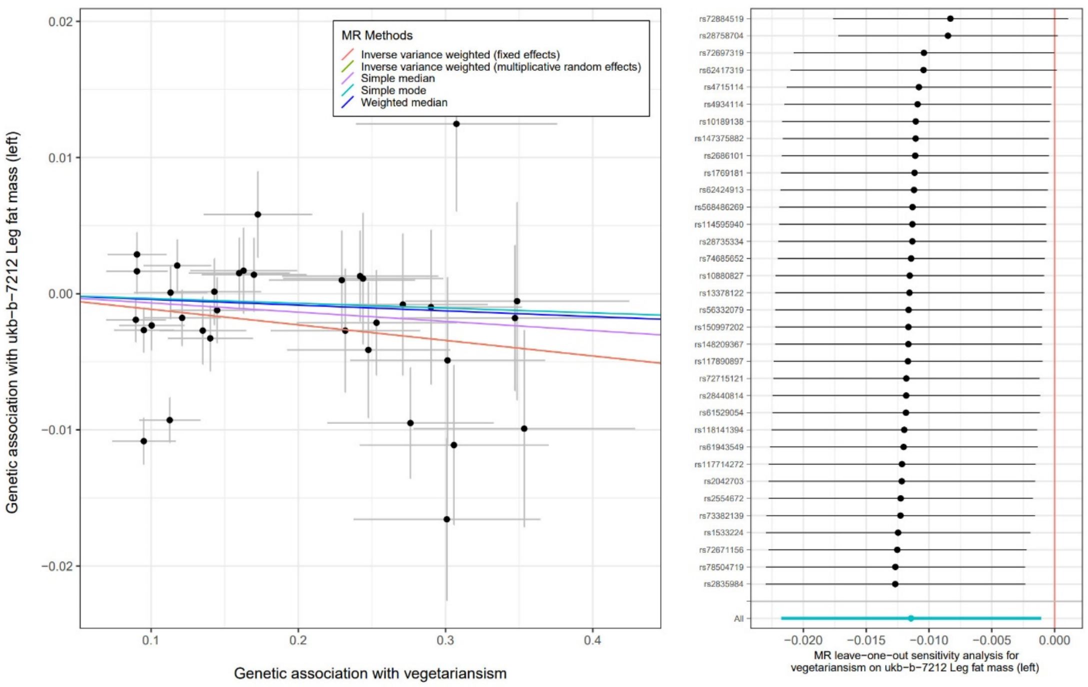


**Supplementary Fig. 26.** Scatter plot for the causal association between vegetarianism and leg fat mass (left) and leave-one-out tests

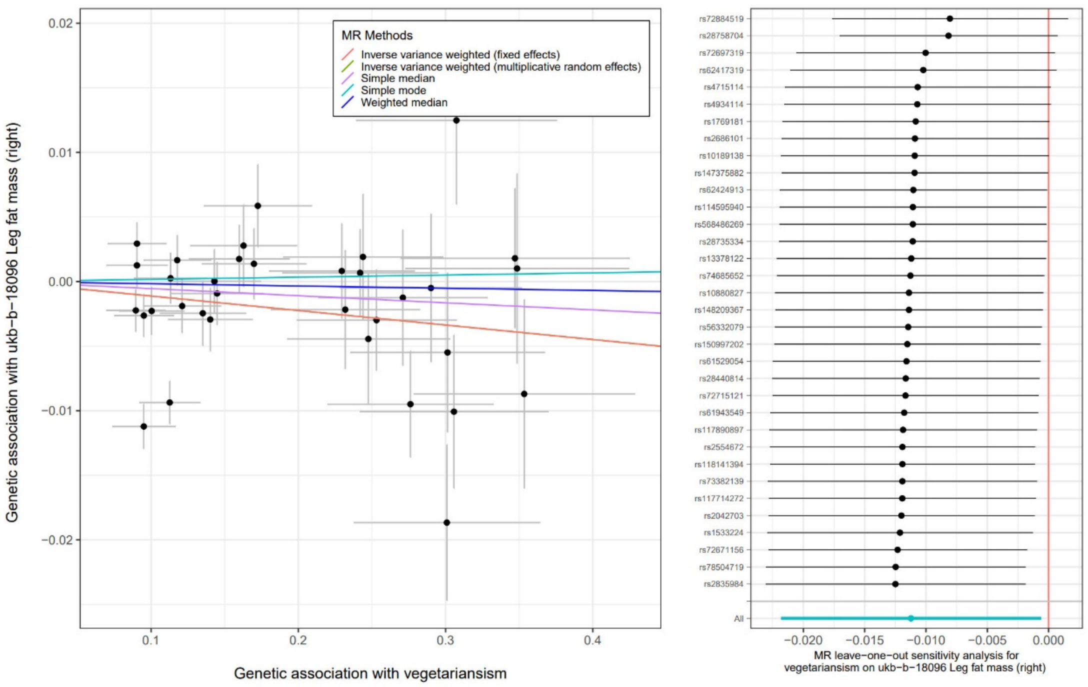


**Supplementary Fig. 27.** Scatter plot for the causal association between vegetarianism and leg fat mass (right) and leave-one-out tests

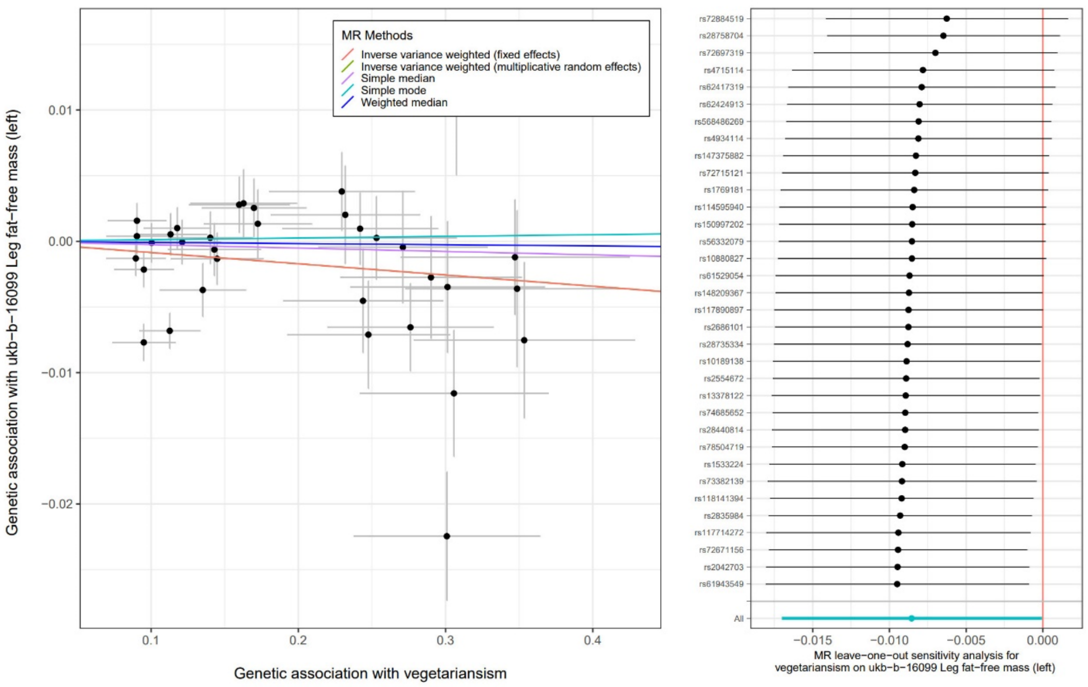


**Supplementary Fig. 28.** Scatter plot for the causal association between vegetarianism and leg fat-free mass (left) and leave-one-out tests

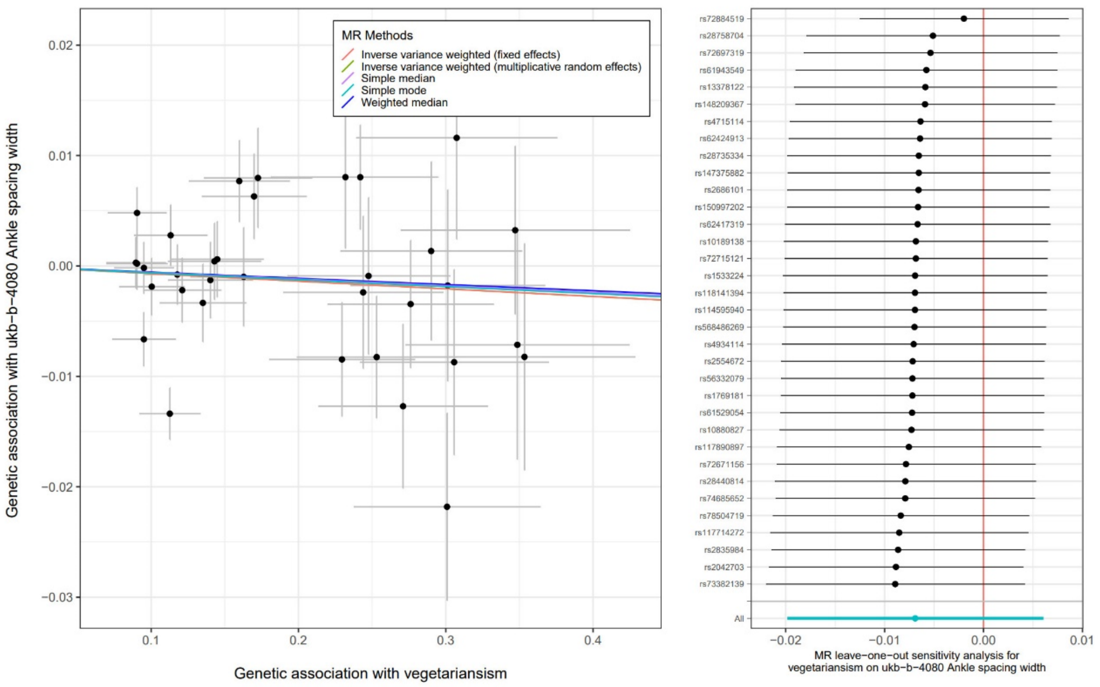


**Supplementary Fig. 29.** Scatter plot for the causal association between vegetarianism and ankle spacing width and leave-one-out tests

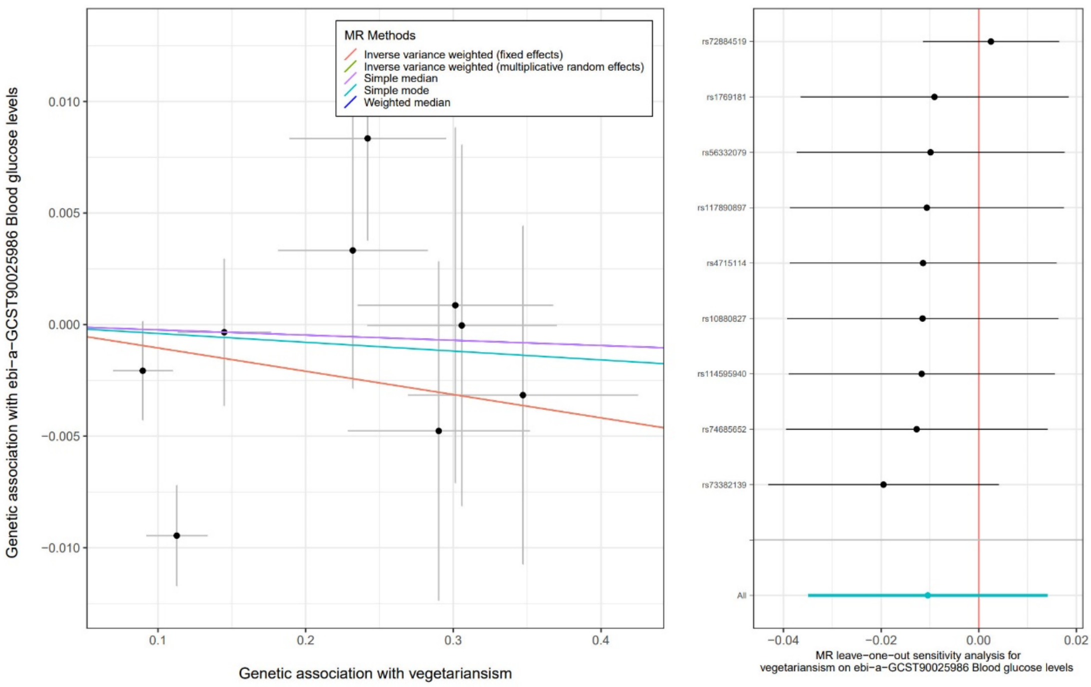


**Supplementary Fig. 30.** Scatter plot for the causal association between vegetarianism and blood glucose levels and leave-one-out tests

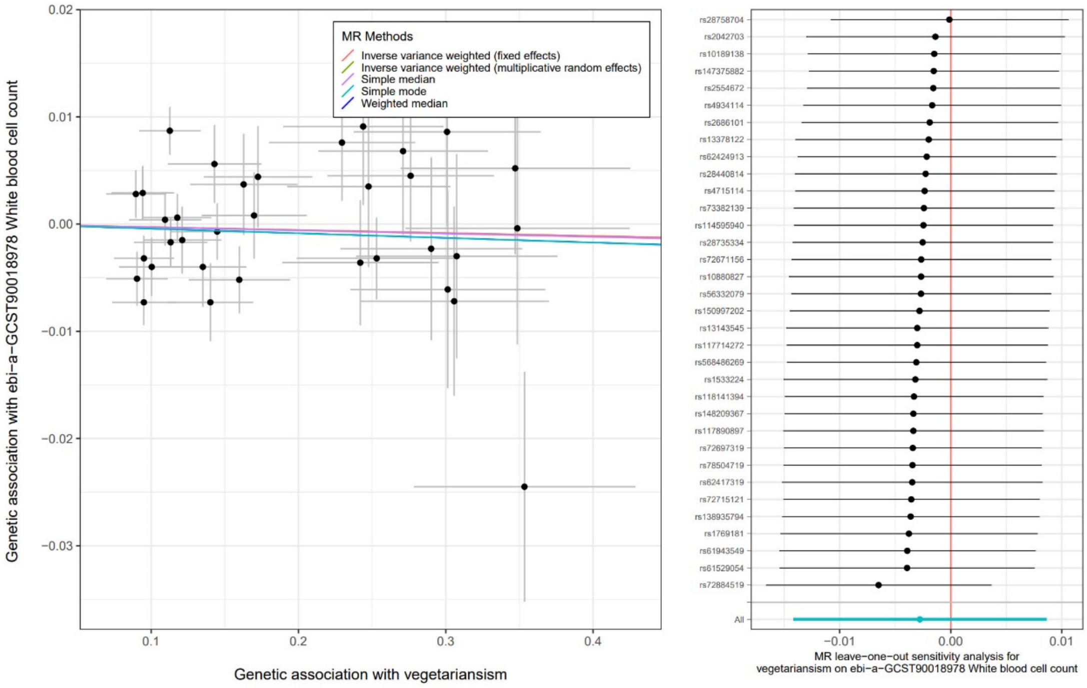


**Supplementary Fig. 31.** Scatter plot for the causal association between vegetarianism and white blood cell count and leave-one-out tests

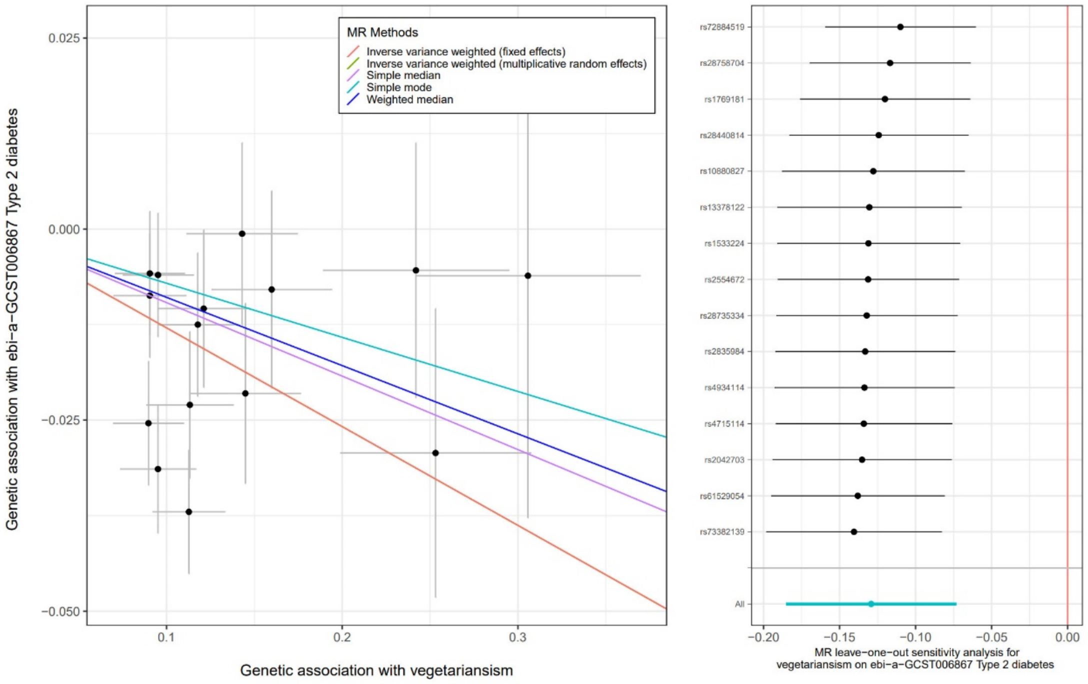


**Supplementary Fig. 32.** Scatter plot for the causal association between vegetarianism and type 2 diabetes and leave-one-out tests

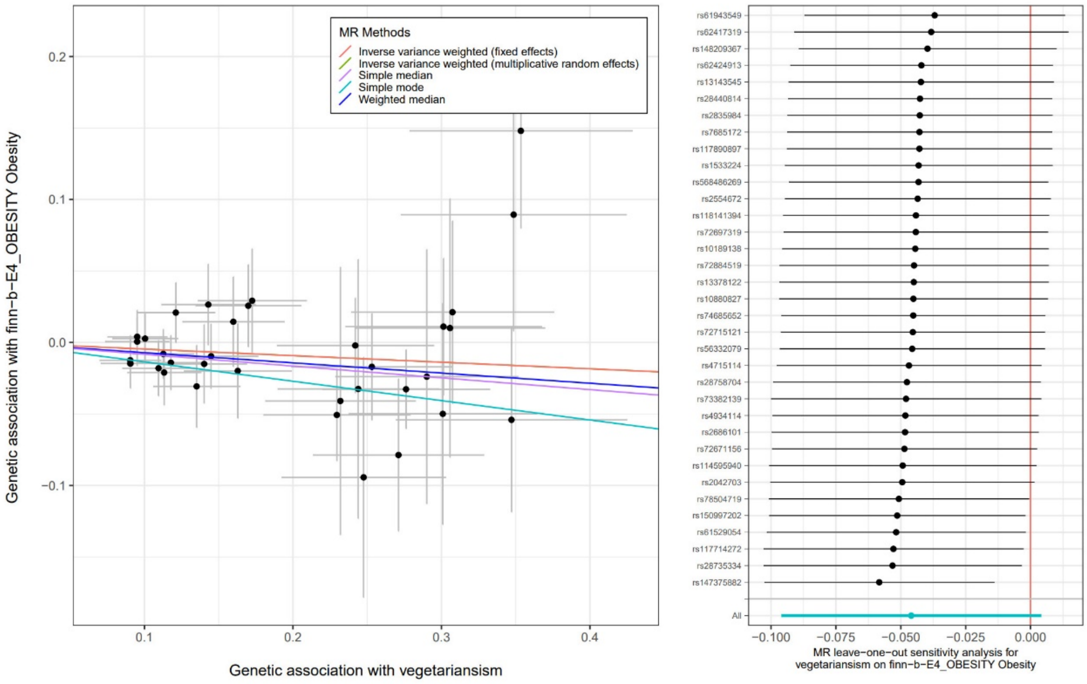


**Supplementary Fig. 33.** Scatter plot for the causal association between vegetarianism and obesity and leave-one-out tests

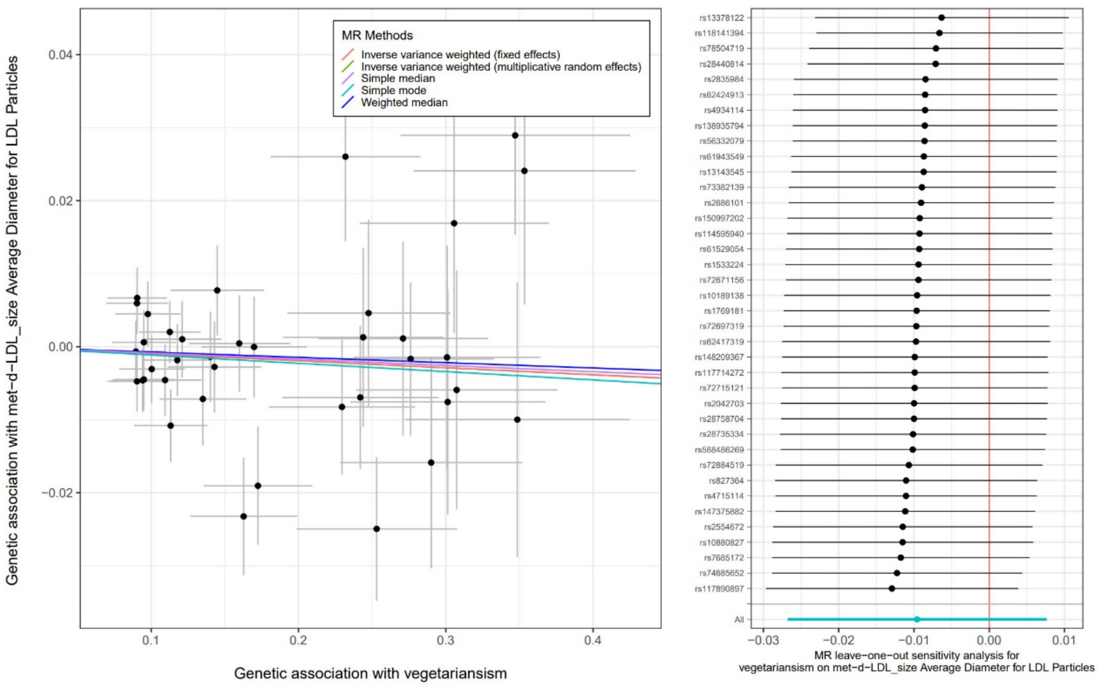


**Supplementary Fig. 34.** Scatter plot for the causal association between vegetarianism and average diameter for LDL particles and leave-one-out tests

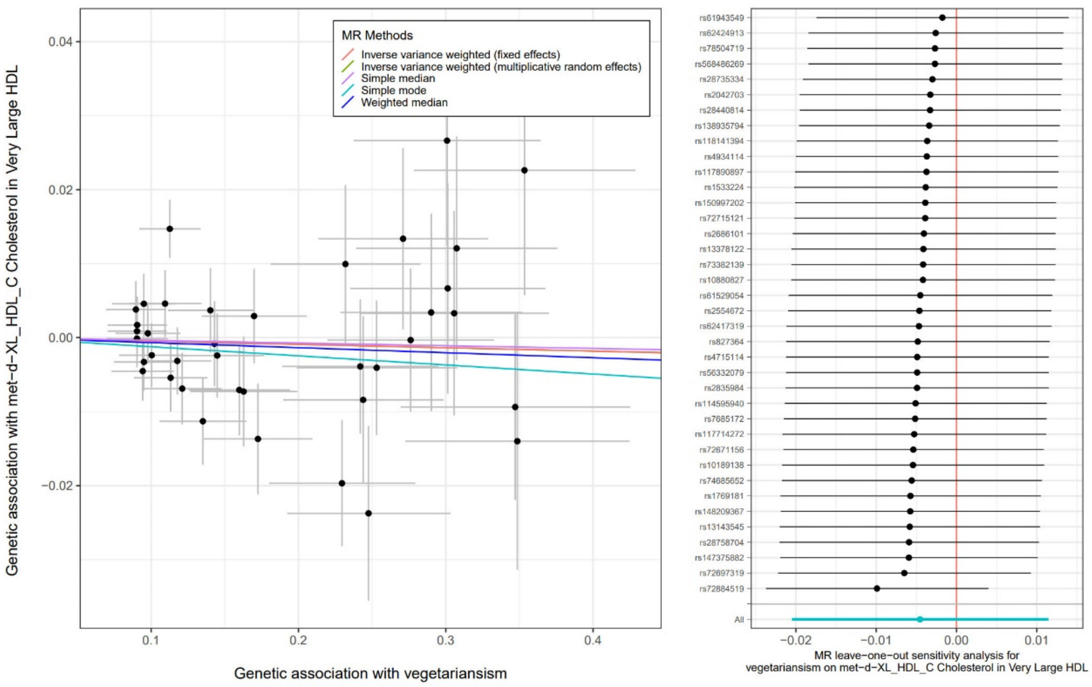


**Supplementary Fig. 35.** Scatter plot for the causal association between vegetarianism and cholesterol in very large HDL and leave-one-out tests

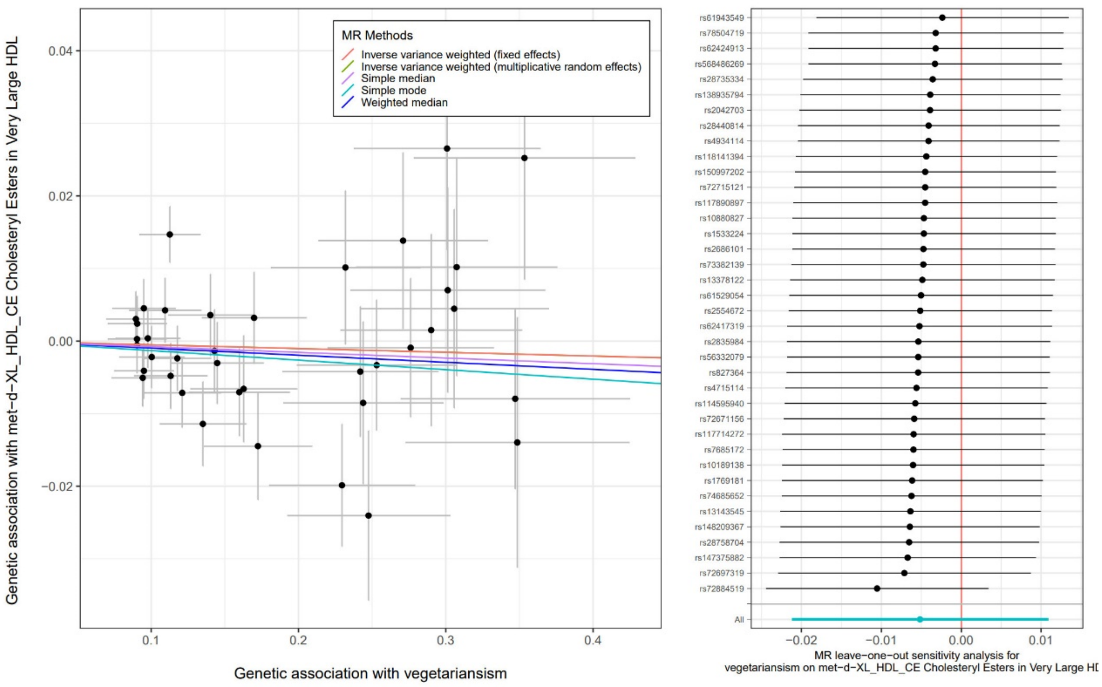


**Supplementary Fig. 36.** Scatter plot for the causal association between vegetarianism and cholesteryl esters in very large HDL and leave-one-out tests

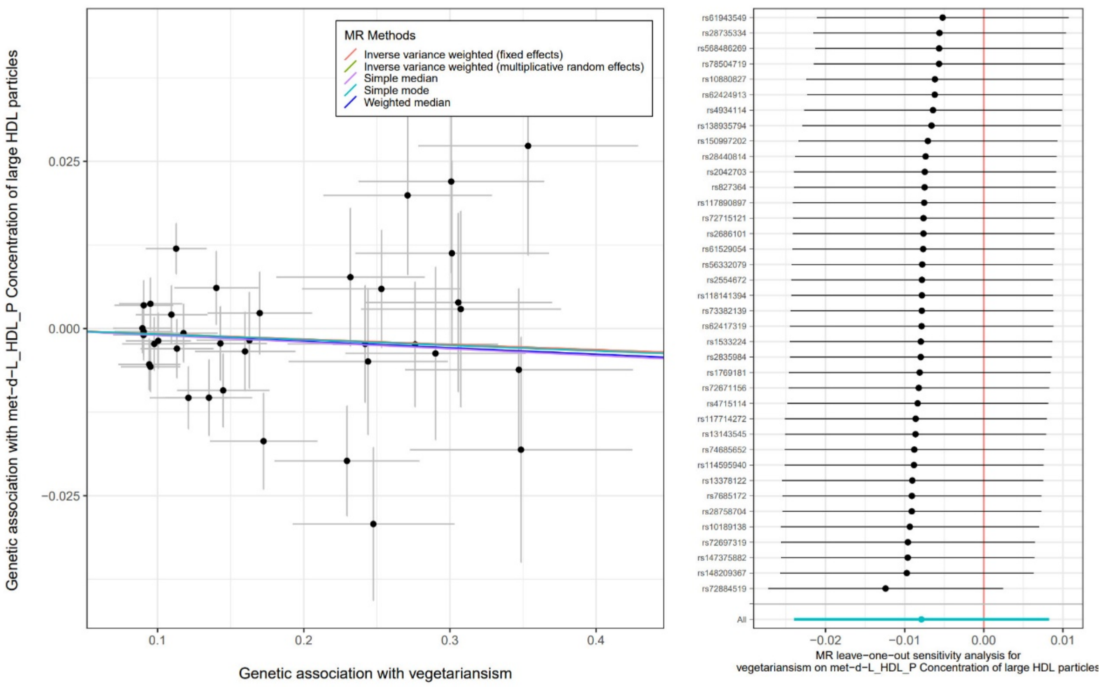


**Supplementary Fig. 37.** Scatter plot for the causal association between vegetarianism and concentration of large HDL particles and leave-one-out tests

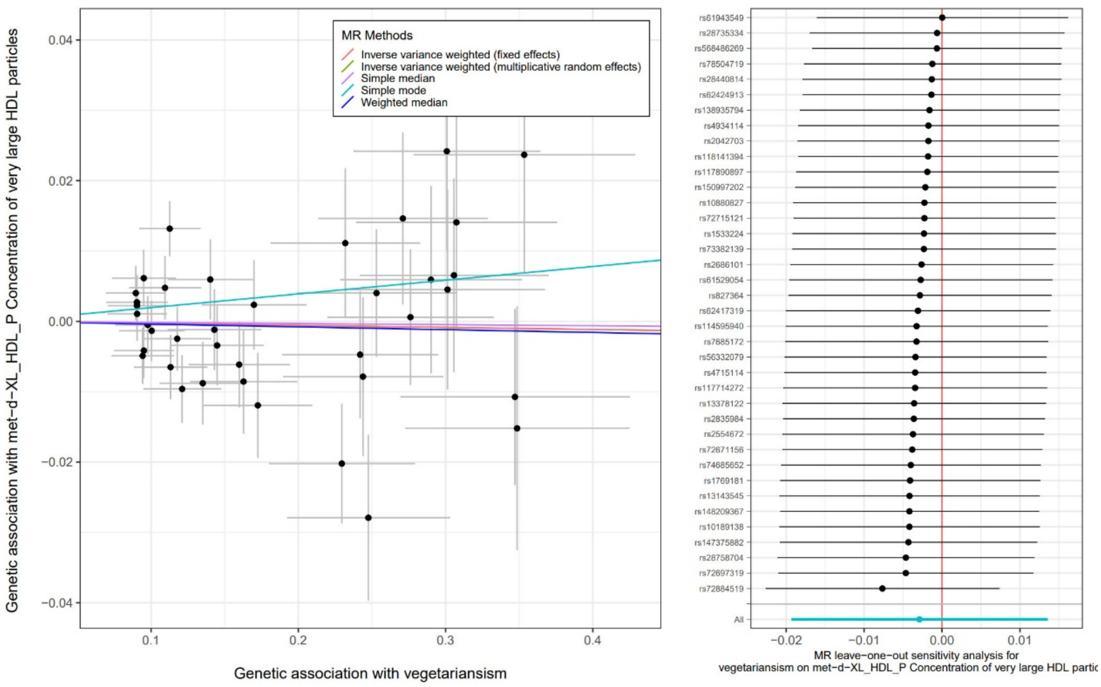


**Supplementary Fig. 38.** Scatter plot for the causal association between vegetarianism and concentration of very large HDL particles and leave-one-out tests

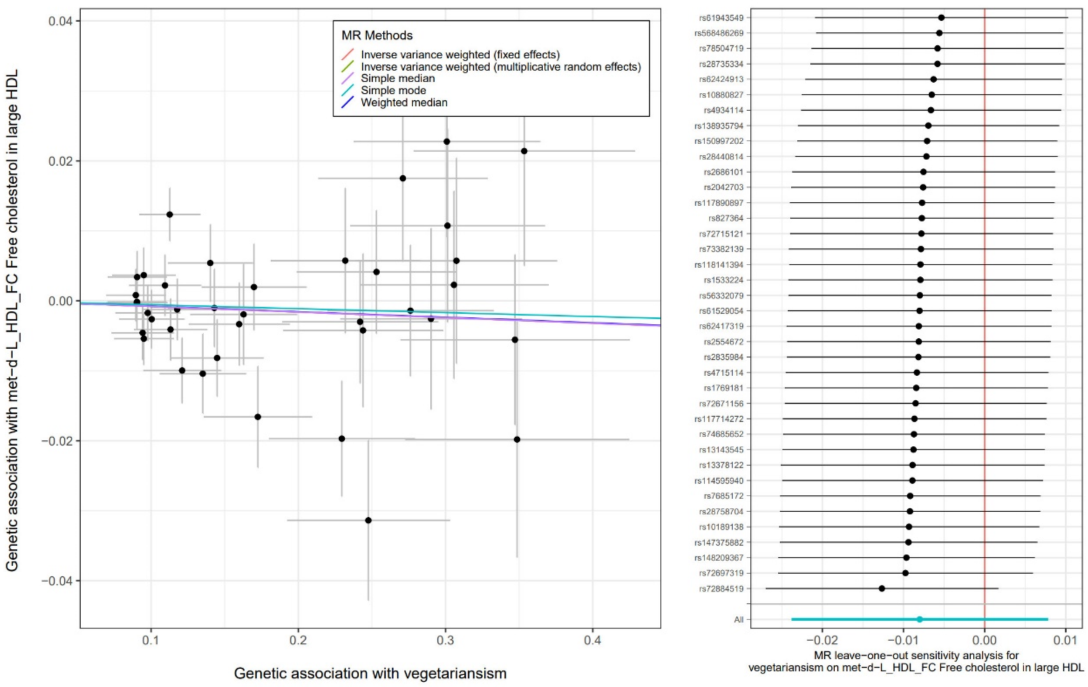


**Supplementary Fig. 39.** Scatter plot for the causal association between vegetarianism and free cholesterol in large HDL and leave-one-out tests

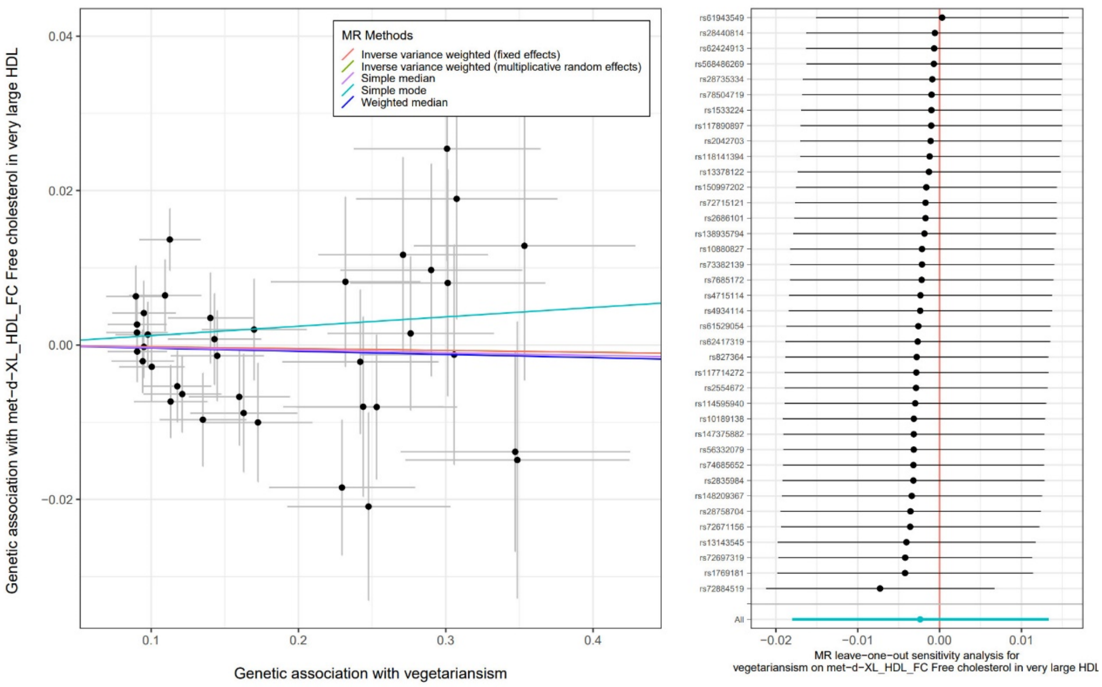


**Supplementary Fig. 40.** Scatter plot for the causal association between vegetarianism and free cholesterol in very large HDL and leave-one-out tests

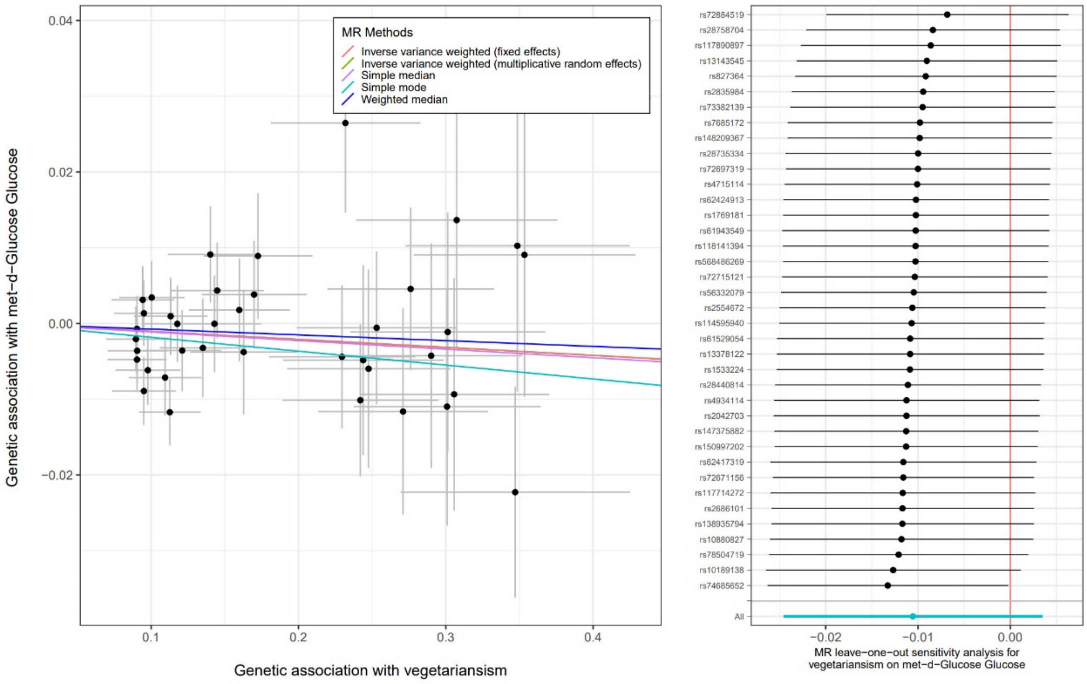


**Supplementary Fig. 41.** Scatter plot for the causal association between vegetarianism and NMR glucose and leave-one-out tests

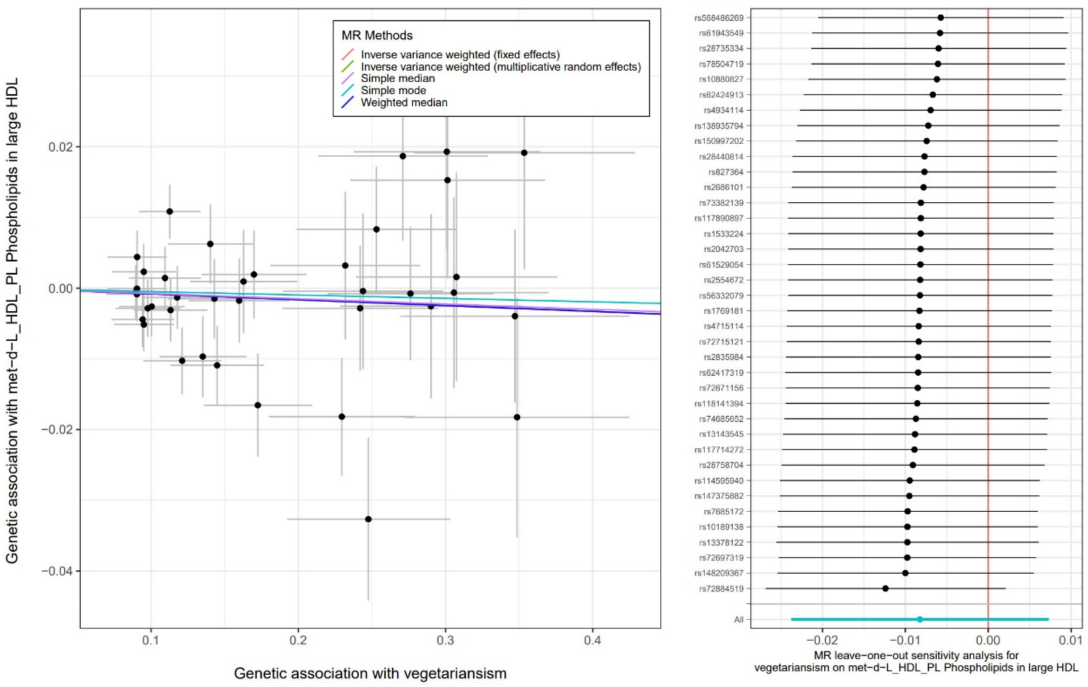


**Supplementary Fig. 42.** Scatter plot for the causal association between vegetarianism and phospholipids in large HDL and leave-one-out tests

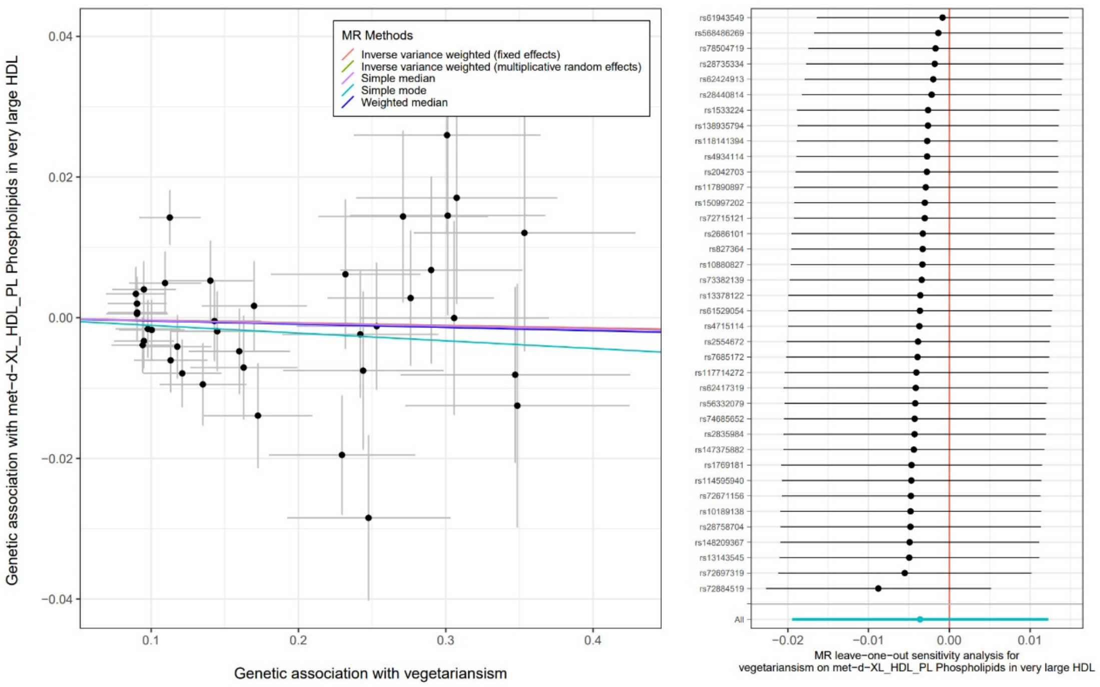


**Supplementary Fig. 43.** Scatter plot for the causal association between vegetarianism and phospholipids in very large HDL and leave-one-out tests

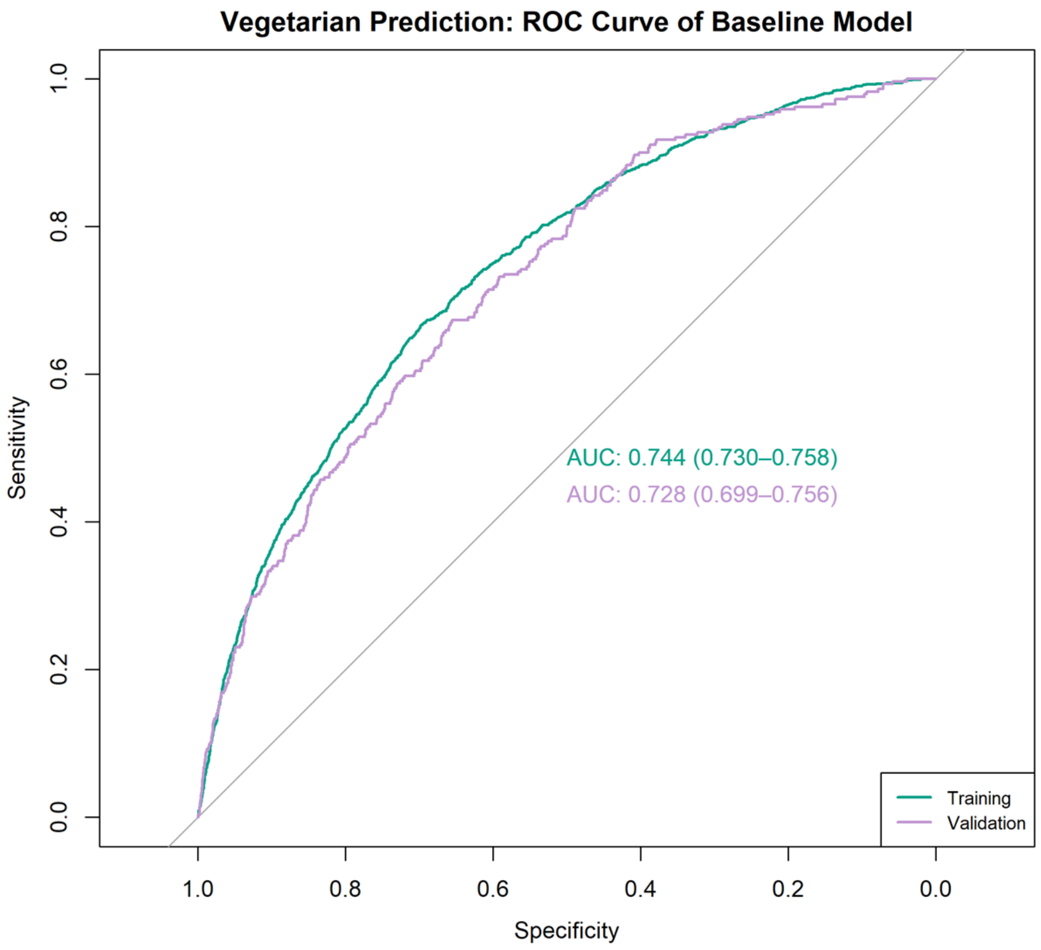


**Supplementary Fig. 44.** AUC plots for the ROC curves of the baseline classification model

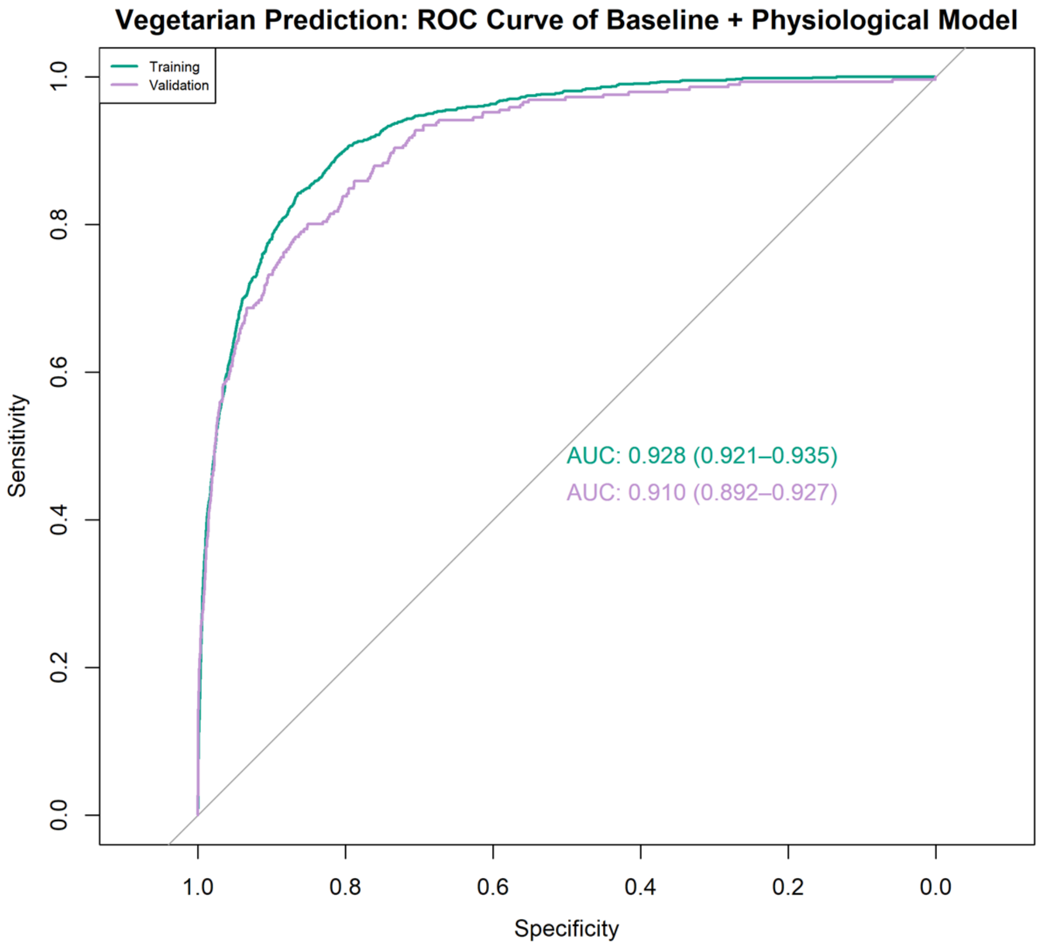


**Supplementary Fig. 45.** AUC plots for the ROC curves of the baseline+genetic classification model

**STROBE-MR checklist of recommended items to address in reports of Mendelian randomization studies**[1 2]

| Item No. | Section | Checklist item | Page No. | Relevant text from manuscript |
|---|---|---|---|---|
| 1 | **TITLE and ABSTRACT** | Indicate Mendelian randomization (MR) as the study's design in the title and/or the abstract if that is a main purpose of the study | 2 | Since MR is not the main purpose of the study, we do not include it in the title, but we do indicate the MR design in the Abstract: "One- and two-sample Mendelian randomization analyses supported causal effects of vegetarianism on 23 outcomes, including lower basal metabolic rate, reduced body mass index (BMI), decreased fat mass, and lower type 2 diabetes risk. " |
| | **INTRODUCTION** | | | |
| 2 | **Background** | Explain the scientific background and rationale for the reported study. What is the exposure? Is a potential causal relationship between exposure and outcome plausible? Justify why MR is a helpful method to address the study question | 4 | "This study aims to address these gaps by systematically investigating a wide array of health, physiological, and behaviour-related outcomes associated with vegetarianism. The goal is to provide a holistic understanding of the implications of plant-based diets, offering insights for nutritionists, policymakers, and individuals considering a transition to vegetarianism. The analysis is grounded in robust causal inference and machine-learning methodologies, addressing limitations of prior observational studies." |
| 3 | **Objectives** | State specific objectives clearly, including pre-specified causal hypotheses (if any). State that MR is a method that, under specific assumptions, intends to estimate causal effects | 4 | "Further, we applied both one-sample and two-sample Mendelian randomization (MR) to investigate potential causal relationships between vegetarianism and the identified health and behaviour-related outcomes." |
| | **METHODS** | | | |
| 4 | **Study design and data sources** | Present key elements of the study design early in the article. Consider including a table listing sources of data for all phases of the study. For each data source contributing to the analysis, describe the following: | | |
| | a) | Setting: Describe the study design and the underlying population, if possible. Describe the setting, locations, and relevant dates, including periods of recruitment, exposure, follow-up, and data collection, when available. | 5 | "Study samples<br>Two independent large cohorts from the UK Biobank (UKB) and the China Health and Nutrition Survey (CHNS) databases were used in the current study. UK Biobank is the database for a population-based study involving more than 500,000 UK residents approved by the NHS National Research |

| | | | | | |
|---|---|---|---|---|---|
| | | | | | UKB sample, including (1) 136 diseases, (2) 10 blood chemistry biomarkers, (3) 44 physical measures, (4) 4 telomere phenotypes, and (5) 197 nuclear magnetic resonance (NMR) metabolomic biomarkers. More details on health and subsequent behaviour-related phenotypes were provided in Supplementary Table 1.<br><br>Behaviour-related phenotypes<br><br>Behaviour-related phenotypes consist of six categories, containing a total of 52 traits: (1) 9 socio-demographic factors (such as education, income, and employment); (2) 25 lifestyle factors (such as alcohol drinking status, social activities, and outdoor activities); (3) 13 mental well-being phenotypes; (4) 5 early-life factors.” |
| | | e) | Provide details of ethics committee approval and participant informed consent, if relevant | 5 | “UK Biobank is the database for a population-based study involving more than 500,000 UK residents approved by the NHS National Research Ethics Service (Ref: 11/NW/0382).” |
| 5 | **Assumptions** | | Explicitly state the three core IV assumptions for the main analysis (relevance, independence and exclusion restriction) as well assumptions for any additional or sensitivity analysis | 8 | “In addition, we conducted several sensitivity analyses to verify the causal relationships identified in one-sample MR analyses. First, a two-sample Mendelian randomization (2SMR) was performed using summary-level data from genome-wide association studies (GWAS). The primary exposure in the 2SMR was vegetarianism (GWAS Catalog ID: GCST90428838), with outcome data sourced from the Medical Research Council Integrative Epidemiology Unit (IEU) OpenGWAS database (https://gwas.mrcieu.ac.uk/). Detailed results are provided in Supplementary Tables 5. Second, Cochran's Q tests were used to evaluate heterogeneity; if heterogeneity was present ($P < 0.05$), a random-effects inverse-variance weighted (IVW-RE) method was employed to address potential bias. Third, we performed MR-Egger intercept tests to assess horizontal pleiotropy. Where pleiotropy was detected ($P < 0.05$), the MR-PRESSO test was used to remove pleiotropic single nucleotide polymorphisms (SNPs), and pleiotropy-corrected results were generated. Lastly, leave-one-out (LOO) analyses were conducted to evaluate the consistency of the combined effect by sequentially removing each SNP, ensuring no |

| | | | | | |
|---|---|---|---|---|---|
| | | | | | individual SNP disproportionately influenced the results." |
| 6 | **Statistical methods: main analysis** | | Describe statistical methods and statistics used | | |
| | | a) | Describe how quantitative variables were handled in the analyses (i.e., scale, units, model) | 6 | "Measurement of vegetarianism<br>A binary variable of vegetarian is defined based on participants' profiles of food intake. Specifically, individuals who did not report any intake of processed meat, beef, pork, lamb/mutton, poultry, fish, seafood and other meat were classified as vegetarians. This resulted in 9,009 vegetarians/486,962 non-vegetarians in the analytical UKB sample, and 2,658 vegetarians/8,984 non-vegetarians in the analytical CHNS sample." |
| | | b) | Describe how genetic variants were handled in the analyses and, if applicable, how their weights were selected | 8 | "Genetic instruments for the exposure were selected from the UKB cohort of 334,779 participants of European ancestry. Instruments were required to meet the following criteria: 1) genome-wide significance (P < 5×10-6); 2) absence of linkage disequilibrium (R2 < 0.01, window size = 10,000 kilobases [kb]); 3) exclusion of potential pleiotropic effects." |
| | | c) | Describe the MR estimator (e.g. two-stage least squares, Wald ratio) and related statistics. Detail the included covariates and, in case of two-sample MR, whether the same covariate set was used for adjustment in the two samples | 8 | "The formal estimation followed a two-stage least squares (2SLS) specification:<br>*First stage:*<br>$Vegetariansism = \delta^{veg}\boldsymbol{GIV} + \mu^{veg}\boldsymbol{X} + \xi^{veg}$ (1)<br>*Second stage:*<br>$Outcome = \gamma^{veg}\widehat{Vegetariansism} + \mu\boldsymbol{X} + \epsilon$ (2)<br>In addition, we conducted several sensitivity analyses to verify the causal relationships identified in one-sample MR analyses. First, a two-sample Mendelian randomization (2SMR) was performed using summary-level data from genome-wide association studies (GWAS). The primary exposure in the 2SMR was vegetarianism (GWAS Catalog ID: GCST90428838), with outcome data sourced from the Medical Research Council Integrative |

| | | | | | |
|---|---|---|---|---|---|
| | | | | | Epidemiology Unit (IEU) OpenGWAS database (https://gwas.mrcieu.ac.uk/). Detailed results are provided in Supplementary Tables 5.” |
| | | d) | Explain how missing data were addressed | 9 | “In each regression, we excluded those with missing data on particular phenotypes, resulting in 28,582 to 458,709 participants in different regressions (Table 1).” |
| | | e) | If applicable, indicate how multiple testing was addressed | 8 | “Post hoc Sidak-Holm corrections were applied to all MR regressions to account for multiple comparisons.” |
| 7 | **Assessment of assumptions** | | Describe any methods or prior knowledge used to assess the assumptions or justify their validity | 16 | “After accounting for heterogeneity and pleiotropy, 23 associations remained significant, confirming robust causal relationships (Fig. 3). All robust associations pertained to health/physiological phenotypes. Detailed estimation results from the random-effects IVW method (robust to the presence of heterogeneity), heterogeneity tests with Cochran’s Q statistics, and MR-Egger intercept tests for horizontal pleiotropy were presented in Supplementary Tables 5. Scatter plots and LOO plots were presented in Supplementary Figures 1-43.” |
| 8 | **Sensitivity analyses and additional analyses** | | Describe any sensitivity analyses or additional analyses performed (e.g. comparison of effect estimates from different approaches, independent replication, bias analytic techniques, validation of instruments, simulations) | 9 | “In addition, we conducted several sensitivity analyses to verify the causal relationships identified in one-sample MR analyses. First, a two-sample Mendelian randomization (2SMR) was performed using summary-level data from genome-wide association studies (GWAS). The primary exposure in the 2SMR was vegetarianism (GWAS Catalog ID: GCST90428838), with outcome data sourced from the Medical Research Council Integrative Epidemiology Unit (IEU) OpenGWAS database (https://gwas.mrcieu.ac.uk/). Detailed results are provided in Supplementary Tables 5. Second, Cochran's Q tests were used to evaluate heterogeneity; if heterogeneity was present ($P < 0.05$), a random-effects inverse-variance weighted (IVW-RE) method was employed to address potential bias. Third, we performed MR-Egger intercept tests to assess horizontal pleiotropy. Where pleiotropy was detected ($P < 0.05$), the MR-PRESSO test was used to remove pleiotropic single nucleotide polymorphisms (SNPs), and pleiotropy-corrected results were generated. Lastly, |

| | | | | | |
|---|---|---|---|---|---|
| | | | | | leave-one-out (LOO) analyses were conducted to evaluate the consistency of the combined effect by sequentially removing each SNP, ensuring no individual SNP disproportionately influenced the results.” |
| 9 | **Software and pre-registration** | | | | |
| | | a) | Name statistical software and package(s), including version and settings used | 7 | “All analyses were performed using R (version 4.4.1).” |
| | | b) | State whether the study protocol and details were pre-registered (as well as when and where) | | The current study protocol was not pre-registered. |
| | **RESULTS** | | | | |
| 10 | **Descriptive data** | | | | |
| | | a) | Report the numbers of individuals at each stage of included studies and reasons for exclusion. Consider use of a flow diagram | 9 | “In each regression, we excluded those with missing data on particular phenotypes, resulting in 28,582 to 458,709 participants in different regressions (Table 1).” |
| | | b) | Report summary statistics for phenotypic exposure(s), outcome(s), and other relevant variables (e.g. means, SDs, proportions) | 15 | Table 2 |
| | | c) | If the data sources include meta-analyses of previous studies, provide the assessments of heterogeneity across these studies | | N/A |
| | | d) | For two-sample MR:<br>i. Provide justification of the similarity of the genetic variant-exposure associations between the exposure and outcome samples<br>ii. Provide information on the number of individuals who overlap between the exposure and outcome studies | 9 | “In addition, we conducted several sensitivity analyses to verify the causal relationships identified in one-sample MR analyses. First, a two-sample Mendelian randomization (2SMR) was performed using summary-level data from genome-wide association studies (GWAS). The primary exposure in the 2SMR was vegetarianism (GWAS Catalog ID: GCST90428838), with outcome data sourced from the Medical Research Council Integrative Epidemiology Unit (IEU) OpenGWAS database (https://gwas.mrcieu.ac.uk/). Detailed results are provided in Supplementary Tables 5. Second, Cochran's Q tests were used to evaluate heterogeneity; if heterogeneity was present (P < 0.05), a random-effects inverse-variance weighted (IVW-RE) method was employed to address potential bias. Third, we performed MR-Egger intercept tests to assess horizontal pleiotropy. |

| | | | | | |
|---|---|---|---|---|---|
| | | | | | suggesting prior inconsistency associations were likely spurious due to endogeneity bias.[22] This comprehensive approach, integrating one-sample and two-sample MR analyses, alongside rigorous sensitivity tests, strengthens the robustness of the findings while acknowledging the complexity of health outcomes linked to vegetarianism.” |
| | | b) | Report MR estimates of the relationship between exposure and outcome, and the measures of uncertainty from the MR analysis, on an interpretable scale, such as odds ratio or relative risk per SD difference | 17 | Fig. 3, Supplementary Tables 4 and 5. |
| | | c) | If relevant, consider translating estimates of relative risk into absolute risk for a meaningful time period | | N/A |
| | | d) | Consider plots to visualize results (e.g. forest plot, scatterplot of associations between genetic variants and outcome versus between genetic variants and exposure) | | N/A |
| 12 | **Assessment of assumptions** | | | | |
| | | a) | Report the assessment of the validity of the assumptions | 16 | “All robust associations pertained to health/physiological phenotypes. Detailed estimation results from the random-effects IVW method (robust to the presence of heterogeneity), heterogeneity tests with Cochran’s Q statistics, and MR-Egger intercept tests for horizontal pleiotropy were presented in Supplementary Tables 5. Scatter plots and LOO plots were presented in Supplementary Figures 1-43.” |
| | | b) | Report any additional statistics (e.g., assessments of heterogeneity across genetic variants, such as $I^2$, Q statistic or E-value) | 16 | Supplementary Table 5, Supplementary Figures 1-43. |
| 13 | **Sensitivity analyses and additional analyses** | | | | |
| | | a) | Report any sensitivity analyses to assess the robustness of the main results to violations of the assumptions | 16 | “All robust associations pertained to health/physiological phenotypes. Detailed estimation results from the random-effects IVW method (robust to the presence of heterogeneity), heterogeneity tests with Cochran’s Q statistics, and MR-Egger intercept tests for horizontal pleiotropy were presented in Supplementary Tables 5. Scatter plots and LOO plots were presented in Supplementary Figures 1-43.” |

| | | | | | |
|---|---|---|---|---|---|
| | | e) | Consider additional plots to visualize results (e.g., leave-one-out analyses) | | Supplementary Figures 1-43. |
| | **DISCUSSION** | | | | |
| 14 | **Key results** | | Summarize key results with reference to study objectives | 20 | "The results of this study highlight the multifaceted health and behavioural impacts of vegetarianism, providing valuable insights into both the positive and nuanced consequences of adopting this dietary pattern. One of the key findings is the robust protective effect of vegetarianism against non-insulin-dependent diabetes mellitus, lower BMI, and reduced fat mass across multiple body regions. These results corroborate prior observational studies that have highlighted the health benefits of plant-based diets in reducing metabolic risks. The causal evidence provided by MR analyses and replications in East Asian populations further strengthens these associations, moving beyond mere correlation to suggest that vegetarianism actively contributes to better metabolic health outcomes across different ethnic groups. In the exploration of using machine-learning models to predict vegetarianism, the inclusion of physiological and genetic factors significantly improved model accuracy, implying that vegetarianism may not merely a lifestyle choice, but one influenced by complex biological mechanisms." |
| 15 | **Limitations** | | Discuss limitations of the study, taking into account the validity of the IV assumptions, other sources of potential bias, and imprecision. Discuss both direction and magnitude of any potential bias and any efforts to address them | 22 | "However, some limitations should be acknowledged. While our MR approach strengthens causal inference, the lack of significant associations with certain traits (e.g., cancer risk) suggests that the protective effects of vegetarianism may be more limited than previously assumed. Additionally, our models are based on UK Biobank data, and though we replicated key findings in an East Asian population (CHNS), further validation across diverse populations is necessary to enhance the generalizability of these results." |
| 16 | **Interpretation** | | | | |
| | | a) | Meaning: Give a cautious overall interpretation of results in the context of their limitations and in comparison with other studies | 21 | "Our findings on the protective effect of vegetarianism against type 2 diabetes align with prior research,[24-25] which consistently links |

| | | | | |
|---|---|---|---|---|
| | | | | plant-based diets to reduced diabetes risk due to their high fibre and low saturated fat content. This study adds causal and cross-population evidence to these observations, reinforcing the benefits of vegetarianism for metabolic health.<br>The causal association with lower basal metabolic rate (BMR) is intriguing, suggesting that vegetarianism may contribute to energy adaptation by reducing caloric needs. While a lower BMR implies fewer calories are required for essential functions,[26] it also presents challenges for individuals transitioning to vegetarianism, potentially necessitating personalized nutritional guidance to avoid energy imbalances.<br>Notably, our results did not find causal links between vegetarianism and cancer or cardiovascular disease risks, challenging earlier studies that suggested protective effects against conditions such as colorectal and breast cancers.[4-5,7,27] This discrepancy may stem from the limitations of observational studies, where confounding variables were not adequately addressed, as highlighted by the inconsistencies in glucose and lipid metabolism results. The use of MR allowed us to mitigate these confounding effects, revealing that previous associations with vegetarianism and metabolic markers may have been biased due to endogeneity.<br>Similarly, while previous research has reported varying associations between vegetarianism and mental health[8,28]—some indicating increased risks of depression and anxiety—our study did not find significant causal links between vegetarianism and mental health outcomes. This discrepancy may reflect the complexity of dietary patterns' psychological impacts, warranting further investigation." |
| | b) | Mechanism: Discuss underlying biological mechanisms that could drive a potential causal relationship between the investigated exposure and the outcome, and whether the gene-environment equivalence assumption is reasonable. Use causal language carefully, clarifying that IV estimates may provide causal effects only under certain assumptions | | NA |
| | c) | Clinical relevance: Discuss whether the results have clinical or public policy relevance, and to what extent they inform effect sizes of possible interventions | 22 | "In conclusion, this study advances our understanding of the health and behavioural |

| | | | | |
|---|---|---|---|---|
| | | | | impacts of vegetarianism by integrating health, physiological, behavioural, and genetic data. Future research should continue to explore the biological underpinnings of dietary behaviour while expanding the scope to include diverse populations and more nuanced health and behavioural outcomes.” |
| 17 | **Generalizability** | Discuss the generalizability of the study results (a) to other populations, (b) across other exposure periods/timings, and (c) across other levels of exposure | 21 | “The causal evidence provided by MR analyses and replications in East Asian populations further strengthens these associations, moving beyond mere correlation to suggest that vegetarianism actively contributes to better metabolic health outcomes across different ethnic groups. ” |
| | **OTHER INFORMATION** | | | |
| 18 | **Funding** | Describe sources of funding and the role of funders in the present study and, if applicable, sources of funding for the databases and original study or studies on which the present study is based | | “This research was funded by the National Natural Science Foundation of China (Grant No. 72103187) and the 2115 Talent Development Program of China Agricultural University.” |
| 19 | **Data and data sharing** | Provide the data used to perform all analyses or report where and how the data can be accessed, and reference these sources in the article. Provide the statistical code needed to reproduce the results in the article, or report whether the code is publicly accessible and if so, where | 9 | “The primary exposure in the 2SMR was vegetarianism (GWAS Catalog ID: GCST90428838), with outcome data sourced from the Medical Research Council Integrative Epidemiology Unit (IEU) OpenGWAS database (https://gwas.mrcieu.ac.uk/). Detailed results are provided in Supplementary Tables 5.” |
| 20 | **Conflicts of Interest** | All authors should declare all potential conflicts of interest | | The authors declare no conflicts of interest. |

This checklist is copyrighted by the Equator Network under the Creative Commons Attribution 3.0 Unported (CC BY 3.0) license.

1. Skrivankova VW, Richmond RC, Woolf BAR, Yarmolinsky J, Davies NM, Swanson SA, et al. Strengthening the Reporting of Observational Studies in Epidemiology using Mendelian Randomization (STROBE-MR) Statement. JAMA. 2021;under review.

2. Skrivankova VW, Richmond RC, Woolf BAR, Davies NM, Swanson SA, VanderWeele TJ, et al. Strengthening the Reporting of Observational Studies in Epidemiology using Mendelian Randomisation (STROBE-MR): Explanation and Elaboration. BMJ. 2021;375:n2233.
